# A multistage, multitask transformer-based framework for multi-disease diagnosis and prediction using personal proteomes

**DOI:** 10.1101/2025.02.19.25322536

**Authors:** Han Li, Yongkang Li, Yukuan Liu, Johnathan Cooper-Knock, Peng Gao, Xiaotao Shen, Shengquan Chen, Xudong Xing, Sai Zhang

## Abstract

Recent advances in cohort-level proteomic profiling have offered unprecedented opportunities for discovering novel biomarkers and developing diagnostic and predictive tools for complex human diseases. However, the inherent complexity of proteomics data and the scarcity of phenotypic labels, particularly for rare diseases, pose significant challenges in modeling proteome-phenome relationships. Utilizing proteomics data from 2,924 plasma proteins measured in 53,014 UK Biobank participants, we introduce Prophet, an interpretable deep learning framework that combines transformer architecture with a multistage, multitask training strategy to improve disease prediction and biological discovery from personal proteomic profiles. Prophet begins with self-supervised pretraining to model protein interactions, followed by prompt-based fine-tuning for disease diagnosis, and concludes with continuous fine-tuning for disease prediction. Extensive benchmarking across more than 100 diseases demonstrates Prophet’s superior performance over multiple baseline methods, achieving the highest increase in the area under the precision-recall curve (AUPRC) by 132.71% for disease diagnosis and 60.29% for disease prediction. Specifically, Prophet enhances diagnostic accuracy for 95.83% of diseases and boosts predictive accuracy for 94.02% of diseases. Through model interpretation, Prophet identifies 21,549 and 25,915 protein-disease associations for prevalent and incident diseases, respectively, and uncovers prevailing proteomics-based similarities among diseases. Our work provides a powerful framework for proteomics-based disease diagnosis, prediction, and biomarker discovery.

## Introduction

Precision medicine is a transformative shift in healthcare, with the aim of tailoring clinical interventions and treatments based on the unique characteristics of individual patients^1–4^. A crucial component of precision medicine is the identification of effective biomarkers, such as circulating proteins, which inform an individual’s health status, disease risk, or treatment response^5–7^. In recent years, the advent of large-scale plasma proteomics databases, such as the UK Biobank Pharma Proteomics Project^8^ (UKB-PPP), provides unparalleled opportunities to discover disease-protein associations, understand disease mechanisms, develop diagnostic and predictive tools, and identify novel therapeutic targets^9–12^. By pairing high-throughput plasma proteomic profiling with detailed phenotypic information, these databases greatly facilitate the discovery of novel biomarkers and the development of effective predictive models, bridging the gap between molecular profiles and clinical outcomes and advancing precision medicine^13–18^.

Modeling the relationship between proteomics and phenotypes presents several challenges. Proteomics datasets typically involve the measurement of thousands of interrelated proteins, which are linked through intricate biological pathways and processes^19–22^. These proteins exhibit non-linear interactions, complicating the identification of their collective effects on disease phenotypes, particularly when using linear methods^14–17^. Additionally, the sparsity of disease labels – due to missing data or low disease prevalence – further complicates the modeling process. The resultant class imbalance, if not properly addressed, can significantly undermine model performance. To overcome these challenges, advanced modeling technologies are needed that are capable of capturing the complex structural relationships intrinsic in proteomics and phenotype data.

In this work, we introduce Prophet (**PRO**teome-**PHE**nome predic**T**or), a transformer-based deep learning framework designed to enhance disease prediction and biological discovery based on proteomics data. Prophet adopts a transformer architecture^23^, along with a multistage, multitask training strategy, to address the aforementioned challenges. In particular, Prophet first leverages self-supervised pretraining to capture genome-wide protein-protein interactions. Based on this pretrained model, Prophet then utilizes continuous prompt-based fine-tuning to predict prevalent diseases for disease diagnosis, and to predict incident diseases for disease prediction, both from personal proteomic profiles. Prophet employs multitasking in which all conditions are predicted at once for each individual and disease relationships are characterized implicitly by introducing a disease cooccurrence loss. Prophet also incorporates an attentive readout module to facilitate the identification of disease-associated proteins.

Through extensive evaluations based on UKB plasma proteomics data, we demonstrated that Prophet significantly outperformed a wide range of baseline methods in both disease diagnosis and prediction. Prophet’s interpretability further enabled systematic identification of disease-associated plasma proteins and disease similarities, yielding not only enhanced predictive accuracy but also valuable biological insights into disease mechanisms. Prophet represents a potent machine learning framework for proteomics-based disease prediction and biological discovery.

## Results

### Overview of Prophet

Proteomics data analyzed in this study were obtained from the UK Biobank Pharma Proteomics Project^8^ (UKB-PPP), which measured 2,924 unique proteins in plasma samples from 53,014 UKB participants. Disease information was extracted from the UKB hospital inpatient records and classified following the FinnGen guidelines^24^. Based on the timing of disease diagnosis relative to the participant’s baseline visit, we defined 120 prevalent and 117 incident diseases (Fig. 1A, Supplementary Table 1; Methods).

**Fig. 1:**
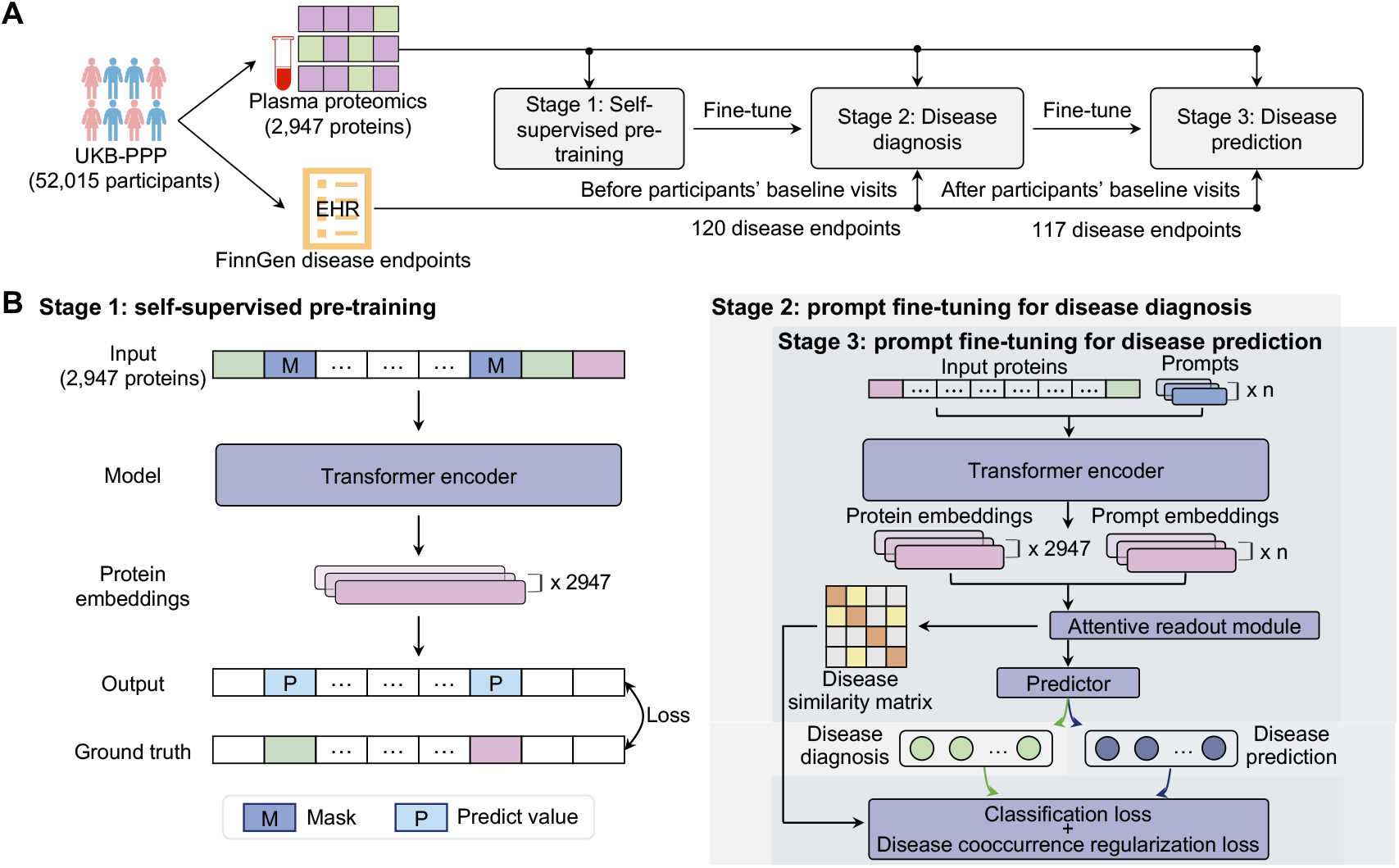
Overview of the Prophet framework for disease diagnosis and prediction using personal proteomics data. (A) Proteomics data analyzed in this study were sourced from UKB-PPP, involving the measurement of 2,924 unique proteins in plasma samples from 53,014 UKB participants. Disease labels were obtained from the hospital inpatient records and classified according to the FinnGen guidelines. A total of 120 prevalent and 117 incident diseases were defined, based on whether they occurred before or after the participant’s baseline visit. The proteomics data, along with the disease labels, were adopted to train Prophet using a multistage, multitask strategy. EHR, electronic health record. (B) With a transformer architecture, Prophet first employs self-supervised pretraining to learn protein-protein interactions. The pretrained model is subsequently fine-tuned using a prompt-based strategy to predict prevalent diseases (i.e., disease diagnosis), followed by additional prompt fine-tuning to predict incident diseases (i.e., disease prediction).

Prophet is a deep learning framework for disease diagnosis and prediction based on personal proteomics data (Fig. 1A; Methods). To capture protein interactions, Prophet first employs a transformer architecture^23^ (Fig. 1B; Methods), a neural network model known for its ability to model long-range dependencies and interactions^25–27^. Additionally, an attentive readout module is introduced to improve model interpretability.

To enhance phenotypic prediction, Prophet adopts a multistage, multitask learning framework (Fig. 1B; Methods). The process begins with self-supervised pretraining by predicting masked protein expression, a technique that has been proven effective in various biological contexts^26, 28–30^. This pretraining strategy allows Prophet to learn generalizable protein representations that capture intricate correlations of protein expression.

Built upon the pretrained model, Prophet next utilizes a prompt-based fine-tuning strategy^31^ to predict prevalent diseases for disease diagnosis (Fig. 1B; Methods). Through this approach, Prophet adapts the network for specific downstream task while retaining valuable knowledge gained from pretraining. Moreover, Prophet implements multitask learning for simultaneous prediction of multiple conditions, aiming to enhance the prediction of diseases with limited labeled data. A disease cooccurrence loss (Fig. 1B; Methods) is particularly introduced here, implicitly encouraging the model to learn disease correlations.

After fine-tuned for prevalent disease prediction, Prophet is continuously fine-tuned for incident disease prediction with the same training strategy (Fig. 1B; Methods). The final trained model can be used for disease diagnosis and prediction for unseen samples, identifying plasma proteins associated with diseases, and uncovering disease similarities based on proteomic signatures.

### Prophet improves disease diagnosis

We first evaluated Prophet’s performance in predicting prevalent diseases (i.e., disease diagnosis). We benchmarked Prophet against six baseline methods, including *k*-nearest neighbors (KNN), Lasso, logistic regression, multilayer perceptron (MLP), random forest, and XGBoost^32^. Both metrics including the area under the receiver operating characteristic (AUROC) and the area under the precision-recall curve (AUPRC) were used for performance evaluation. To assess model robustness, the training and testing procedures were repeated five times with different random seeds for all methods. The dataset splits for training and testing were kept consistent across all models. For 120 prevalent diseases (Methods), Prophet achieved an average AUROC of 0.775 and an average AUPRC of 0.197, with AUROC greater than 0.8 for 44 diseases (36.7%; Supplementary Table 2). Notably, our results demonstrated that Prophet significantly outperformed all baseline methods (*P* = 8.1 *×* 10^−5^, two-sided paired *t*-test, Prophet vs. XGBoost; Fig. 2A), yielding average relative improvements of 4.68% in AUROC and 7.84% in AUPRC (Supplementary Fig. 1A).

**Fig. 2:**
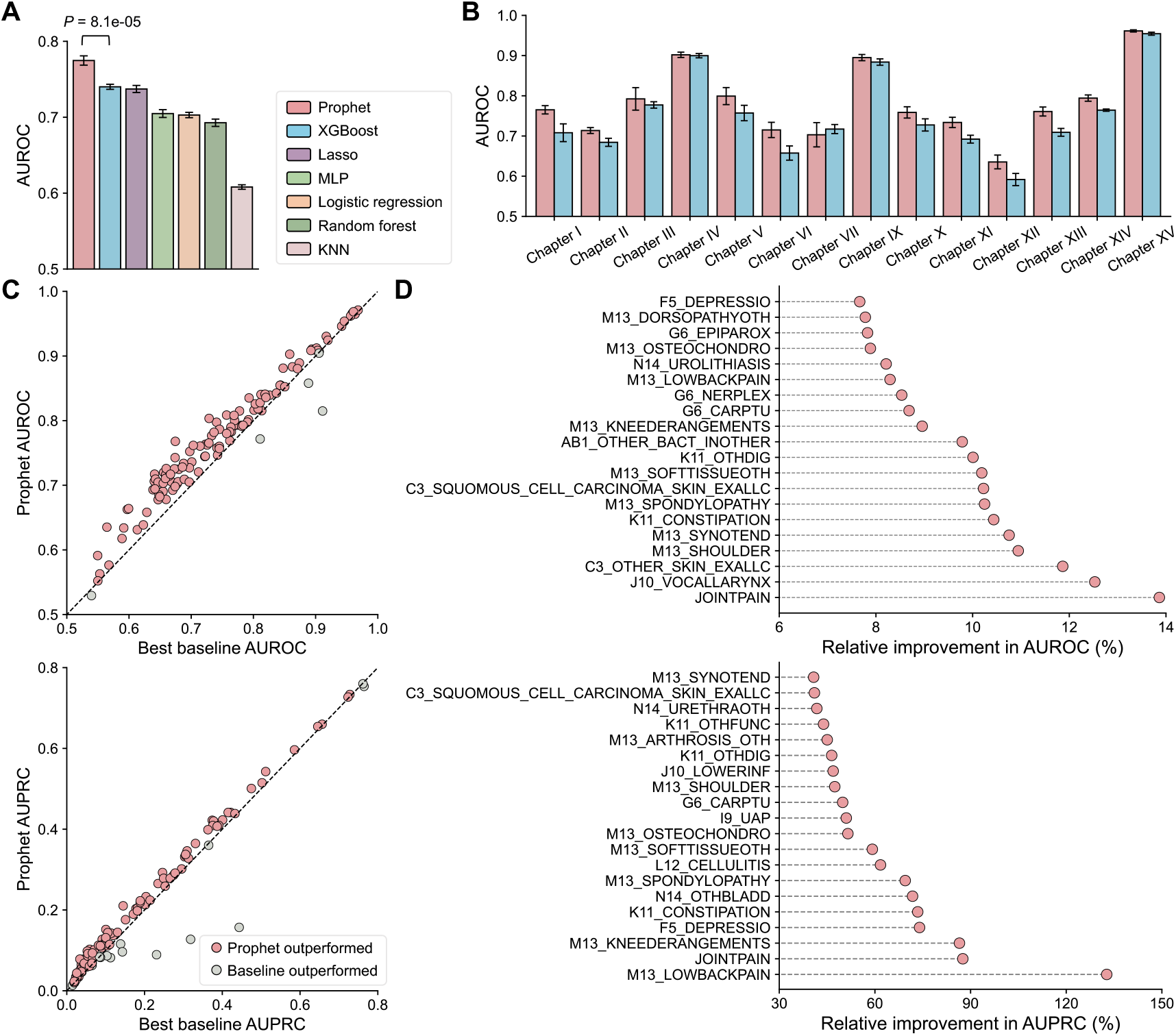
Performance evaluation for disease diagnosis. (A) AUROC scores of Prophet and six baseline methods for predicting 120 prevalent diseases. *P*-value by two-sided paired *t*-test. The bar plot and error bar denote the mean and standard deviation, respectively. AUROC, area under the receiver operating characteristic curve; MLP, multilayer perceptron; KNN, *k*-nearest neighbors. (B) AUROC comparison between Prophet (red) and XGBoost (blue) across different disease categories. (C) Comparison of AUROC (top) and AUPRC (bottom) between Prophet and the best-performing baseline methods. Each dot represents a disease. Red dots denote cases where Prophet outperformed all baseline methods, while gray dots indicate cases where the best baseline method outperformed Prophet. AUPRC, area under the precision-recall curve. (D) Top 20 relative improvements in AUROC (top) and AUPRC (bottom) obtained by Prophet compared to the best-performing baseline methods. The training and testing procedure was conducted for five repeats using different random seeds for all models.

When evaluated across 14 disease categories, Prophet outperformed XGBoost which was the best baseline method on average for 13 categories in AUROC (Fig. 2B), with relative improvements ranging from 0.73% to 8.74%, and for 12 categories in AUPRC (Supplementary Fig. 1B), with relative improvements ranging from 1.55% to 34.04%. In particular, Prophet exhibited markedly superior performance in categories such as Chapter VI: Diseases of the nervous system (8.74% in AUROC and 12.57% in AUPRC), Chapter I: Certain infectious and parasitic diseases (8.06% in AUPRC and 21.8% in AUPRC), Chapter XII: Diseases of the skin and subcutaneous tissue (7.37% in AUROC and 34.04% in AUPRC), Chapter XIII: Diseases of the musculoskeletal system and connective tissue (7.30% in AUROC and 20.72% in AUPRC), Chapter XI: Diseases of the digestive system (6.00% in AUROC and 15.60% in AUPRC), and Chapter V: Mental and behavioural disorders (5.56% in AUROC and 31.90% in AUPRC; Fig. 2B and Supplementary Fig. 1B).

In the analysis of individual diseases, Prophet outperformed the best-performing baseline methods for 95.83% of the 120 diseases (*n* = 115) in AUROC (Fig. 2C). Among these, it achieved a relative improvement of over 10% for 10 diseases and greater than 5% for 45 diseases, with a maximum improvement of 13.86% for JOINTPAIN (Pain in joint). For AUPRC, Prophet surpassed the best-performing baseline models for 88.33% of the 120 diseases (*n* = 106; Fig. 2C), showing a relative improvement of more than 30% for 30 diseases and over 10% for 71 diseases, and with the highest improvement of 132.71% for M13 LOWBACKPAIN (Low back pain). The top 20 relative improvements achieved by Prophet compared to the best baseline methods are shown in Fig. 2D. These results highlight Prophet’s potential as an effective diagnostic tool.

To examine the contribution of different components in Prophet, we conducted extensive ablation studies (Supplementary Fig. 2). Overall, Prophet consistently outperformed its variants (Supplementary Fig. 2A), underscoring the important role of each component in enhancing predictive performance. Interestingly, pretraining provided the upmost improvement, with Prophet achieving a 1.76% relative increase in AUROC compared to the model without pretraining. Additionally, Prophet displayed robust performance across a wide range of prompt numbers, with slight improvement observed as the prompt number increased from one to six (Supplementary Fig. 2B). However, performance plateaued when the prompt number was further increased to 10. We also evaluated the influence of varying disease cooccurrence regularization coefficients in the loss function. Prediction performance improved with increasing coefficients up to 10 (Supplementary Fig. 2C), further emphasizing the importance of this regularization term in optimizing Prophet.

### Prophet enhances disease prediction

Next, we inspected the prediction of incident diseases using Prophet. In particular, for 117 incident diseases, Prophet obtained an average AUROC of 0.718 and an average AUPRC of 0.208, with AUROC greater than 0.75 for 40 diseases (34.2%; Supplementary Table 1). As with disease diagnosis, our comparison showed that Prophet significantly outperformed all baseline methods, achieving average relative improvements of 4.03% in AUROC (Fig. 3A) and 8.50% in AUPRC (Supplementary Fig. 3A) across 117 incident diseases.

**Fig. 3:**
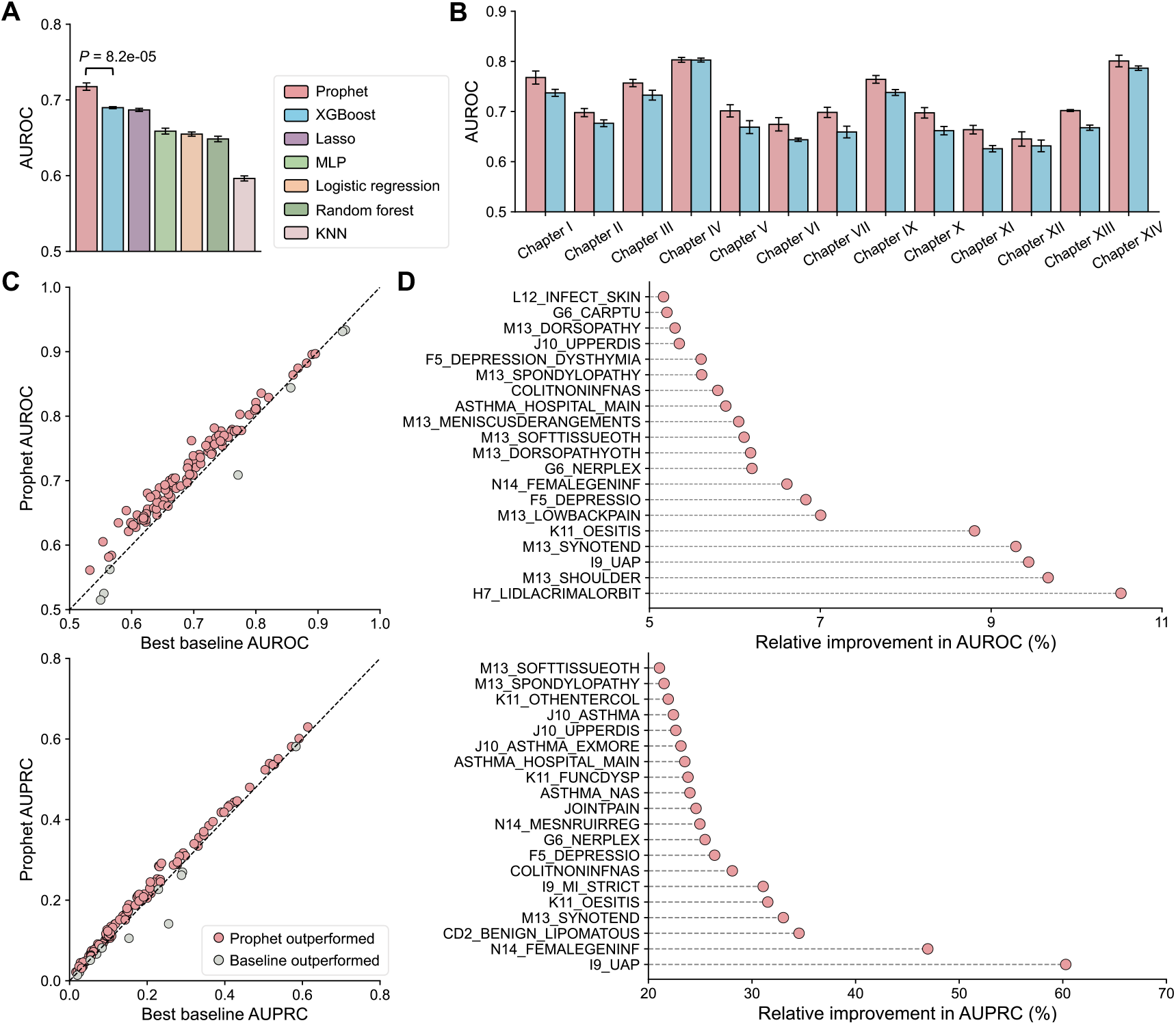
Performance evaluation for disease prediction. (A) AUROC scores of Prophet and six baseline methods for predicting 117 incident diseases. *P*-value by two-sided paired *t*-test. The bar plot and error bar denote the mean and standard deviation, respectively. AUROC, area under the receiver operating characteristic curve; MLP, multilayer perceptron; KNN, *k*-nearest neighbors. (B) AUROC comparison between Prophet (red) and XGBoost (blue) across different disease categories. (C) Comparison of AUROC (top) and AUPRC (bottom) between Prophet and the best-performing baseline methods. Each dot represents a disease. Red dots denote cases where Prophet outperformed all baseline methods, whereas gray dots indicate cases where the best baseline method outperformed Prophet. AUPRC, area under the precision-recall curve. (D) Top 20 relative improvements in AUROC (top) and AUPRC (bottom) obtained by Prophet compared to the best baseline methods. The training and testing procedure was conducted for five repeats using different random seeds for all models.

Notably, Prophet outperformed XGBoost for all 14 disease categories evaluated in AUROC (Fig. 3B), with relative improvements ranging from 0.05% to 6.07%, and for 13 out of 14 categories in AUPRC (Supplementary Fig. 3B), with relative improvements ranging from 4.56% to 16.83%. Prophet displayed considerable gains in several categories, including Chapter XI: Diseases of the digestive system (6.07% in AUROC and 15.07% in AUPRC), Chapter VII: Diseases of the eye, adnexa, ear and mastoid process (5.94% in AUPRC and 5.44% in AUPRC), Chapter X: Diseases of the respiratory system (5.38% in AUROC and 16.83% in AUPRC), Chapter XIII: Diseases of the musculoskeletal system and connective tissue (5.11% in AUROC and 12.14% in AUPRC), and Chapter V: Mental and behavioural disorders (4.85% in AUROC and 10.71% in AUPRC) (Fig. 3B and Supplementary Fig. 3B).

For individual diseases, Prophet outperformed the best-performing baseline methods for 94.02% of the 117 diseases (*n* = 110) in AUROC (Fig. 3C), and it achieved relative improvements of greater than 5% for 24 diseases, with the largest improvement of 10.52% for H7 LIDLACRIMALORBIT (Disorders of eyelid, lacrimal system and orbit). In terms of AUPRC, Prophet excelled the best-performing baseline models for 90.59% of diseases (*n* = 106; Fig. 3C), with the largest relative improvement of 60.29% for I9 UAP (Unstable angina pectoris), and with improvements of over 30% for 6 diseases and more than 10% for 59 diseases. The top 20 relative improvements obtained by Prophet compared to the best baseline methods are shown in Fig. 3D. These findings highlight the potential of Prophet as a disease prediction tool based on proteomic profiles.

### Prophet stratifies disease risk

We further sought to evaluate Prophet’s ability to stratify disease risk under different scenarios. First, we assessed Prophet’s predictive performance over time. For disease prediction, Prophet achieved average AUROC scores of 0.741, 0.710, and 0.688 under time intervals of 5-year, 10-year, and *>*10-year, respectively. Interestingly, both Prophet and XGBoost showed a reduction in performance with longer time intervals, but Prophet consistently outperformed XGBoost at every time window (Fig. 4A). In contrast, disease diagnostic models did not exhibit the same performance reduction over time; however, Prophet still maintained a consistent advantage over XG-Boost across all time intervals (Supplementary Fig. 4A).

**Fig. 4:**
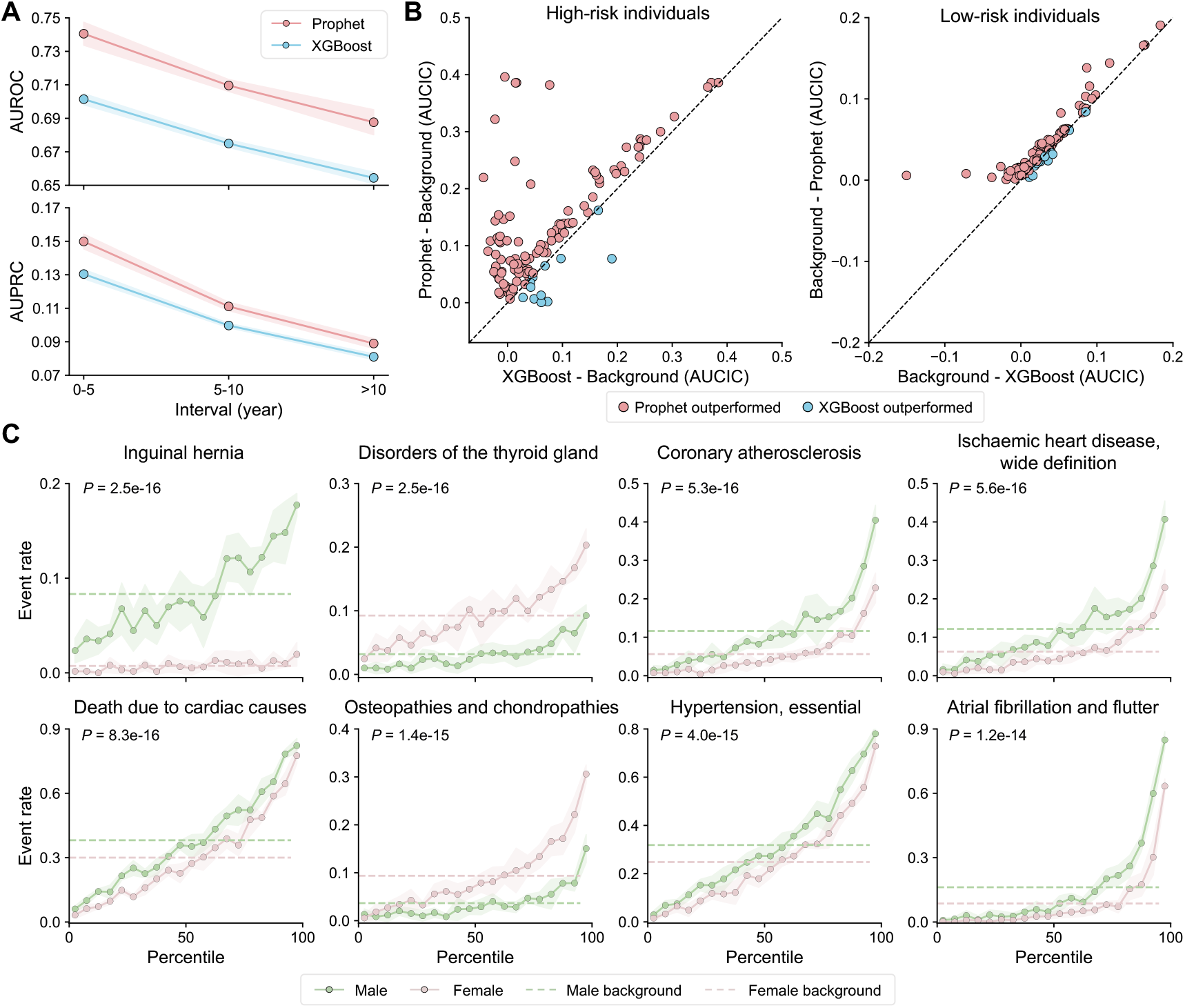
Evaluation of risk stratification based on Prophet for disease prediction. (A) Model performance for disease prediction across different time windows. The positive-to-negative ratio was ensured to be consistent across time windows. The dot and the shaded region denote the mean and standard deviation, respectively. AUROC, area under the receiver operating characteristic curve; AUPRC, area under the precision-recall curve. (B) Comparison of the area under the cumulative incidence curve (AUCIC) between Prophet and XGBoost for high-risk individuals (right) and low-risk individuals (left). High-risk and low-risk individuals are defined as those individuals whose risk scores of incident diseases are in the top 5% and bottom 5% of all predictions, respectively. Each dot represents a disease. (C) Incidence rates for the eight diseases with the largest difference between sexes, against Prophet’s prediction percentiles. *P*-value by two-sided Wilcoxon signed-rank test. The dot and shaded area represent the mean and standard deviation, respectively. The male background (green dashed line) and female background (red dashed line) are defined as the estimated incidence rates of males and females, respectively.

Next, we examined the effectiveness of Prophet in disease risk stratification (Methods). To achieve this, we compared the area under the cumulative incidence curve (AUCIC) between high-risk/low-risk individuals and the background population. High-risk and low-risk individuals were defined based on a specific predictor, with high-risk individuals having prediction scores in the top 5% and low-risk individuals in the bottom 5% of all predictions. A more effective stratification method should yield a higher AUCIC score for high-risk individuals and a lower AUCIC score for low-risk individuals, compared to the background population (Supplementary Fig. 5). Significantly, in the context of disease prediction, Prophet achieved an average AUCIC of 0.180 for its high-risk individuals, surpassing XGBoost by a relative increase of 46.70%. In particular, Prophet yielded higher AUCIC scores than the background for all 117 incident diseases, outperforming XGBoost in 89.74% of the diseases (*n* = 105; Fig. 4B). For its low-risk individuals, Prophet achieved an average AUCIC of 0.011, demonstrating a 34.23% relative reduction compared to XGBoost. Again, Prophet yielded lower AUCIC scores than the background for all 117 diseases, outperforming XGBoost in 86.32% of the diseases (*n* = 101; Fig. 4B). Of note, Prophet yielded a zero AUCIC for 10 diseases, indicating that no low-risk individuals were diagnosed during the follow-up period.

**Fig. 5:**
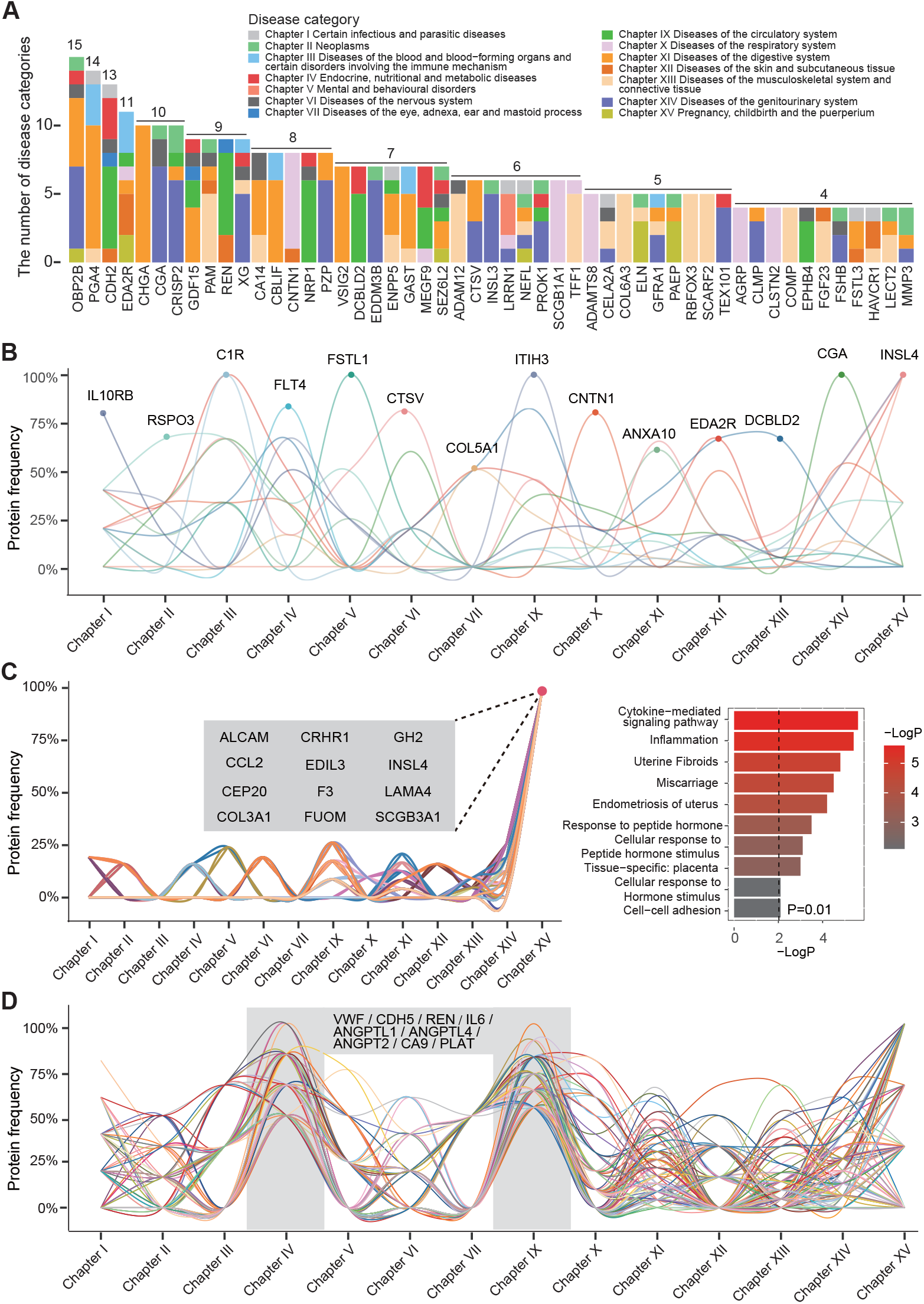
Prophet identifies plasma proteins related to disease diagnosis. (A) Stacked bar chart of protein importance in diagnosing diseases, with colors representing different disease categories (throughout this figure). The number above each bar represents the count of diseases wherein the protein was ranked in the top 5. (B) Line plot showing the frequencies of associated proteins across disease categories. Representative proteins are highlighted with dots and labeled by their names. The frequency of a specific protein in a disease category was defined by the proportion of diseases within that category in which the protein was identified as an associated factor. (C) Functional enrichment analysis of Chapter XV-specific proteins. *P*-value by hypergeometric test. (D) Disease-associated proteins shared between Chapter IV and Chapter IX.

For disease diagnostic models, similar patterns were observed in risk stratification. For high-risk individuals, Prophet achieved an average AUCIC of 0.0846, outperforming XGBoost in 85.83% of 120 prevalent diseases (*n* = 103), with a relative increase of 8.52% (Supplementary Fig. 4B). For low-risk individuals, Prophet obtained an average AUCIC of 0.003, exceeding XGBoost in 72.50% of the diseases (*n* = 87), with a relative decrease of 24.18% (Supplementary Fig. 4B).

Lastly, we investigated the impact of sex on Prophet’s prediction (Methods). With the disease predictive model, 66.67% of the diseases (*n* = 78) exhibited significant sex-dependent difference in risk stratification (adjusted *P <* 0.05, two-sided Wilcoxon signed-rank test; Fig. 4C, Supplementary Table 4). Interestingly, the incidence rate increased progressively with higher prediction percentiles for both sexes, initially below the background rate, but eventually exceeding it. This further highlighted the effectiveness of Prophet for risk stratification. Similarly, the disease diagnostic model uncovered 58.33% of the diseases (*n* = 70) with significant sex difference in stratification (adjusted *P <* 0.05, two-sided Wilcoxon signed-rank test; Supplementary Fig. 4C, Supplementary Table 4).

### Prophet identifies disease-associated plasma proteins

In addition to disease prediction, Prophet also enables the identification of disease-associated proteins through model interpretation. Using attention scores learned by Prophet (Methods), we identified 21,549 protein-disease associations (PDAs) from the diagnostic model, and 25,915 PDAs based on the predictive model, where 11,636 PDAs were shared between these two models (Supplementary Table 5).

Prophet not only successfully pinpointed disease-associated proteins shared across different disease categories (Fig. 5A), such as CHGA, GDF15, and VSIG2, but also demonstrated capacity in identifying disease-specific proteins (Fig. 5B). For instance, IL10RB, an essential protein involved in the antiviral process^33^, was identified as a specific predictor for Chapter I (Certain infectious and parasitic diseases). Additionally, FSTL1, known for its role in the development and diseases of the central nervous system^34^, emerged as a specific biomarker for Chapter V (Mental and behavioral disorders). In the disease category of Chapter XV (Pregnancy, childbirth, and the puerperium), we identified a substantial number of disease-specific proteins (Fig. 5C, left). Functional enrichment analysis revealed that these proteins exhibited consistency with disease phenotypes, being closely associated with functions such as hormonal response and immune regulation (Fig. 5C, right). Notably, Prophet-prioritized proteins for diseases in Chapter IV (Endocrine, nutritional and metabolic diseases) and Chapter IX (Diseases of the circulatory system) displayed a degree of synergy, indicating that these two disease categories share common underlying factors (Fig. 5D). Indeed, previous studies have established a strong association between circulatory diseases and metabolic disorders^35, 36^. This reflects current clinical practice where enhanced screening for cardiovascular disease is recommended for all patients with endocrine disorders^37^. Our findings provided further evidence supporting the overlapping between these two disease groups, exemplified by the shared protein markers such as VWF, CDH5, and ANGPT2, offering new insights into clinical diagnosis and treatment strategies.

We further examined the significant proteins identified by the disease predictive Prophet model (Methods). Overall, disease-associated proteins prioritized by the diagnostic and predictive models exhibited varying degrees of consistency across different disease categories (Fig. 6A and 6B, Supplementary Table 4), indicating that many proteins played both diagnostic and predictive roles. Importantly, EDA2R and GDF15 emerged as the top-ranked predictive biomarkers across a large number of diseases (Fig. 6A), consistent with previous studies^13, 14, 17^. The biological functions of Prophet-identified proteins informed disease mechanisms (Fig. 6C and 6D). For instance, CBLIF (Cobalamin Binding Intrinsic Factor), a glycoprotein essential for Vitamin B12 absorption and erythrocyte maturation^38^, presented a strong association with blood-related diseases (Chapter III) and metabolic diseases (Chapter IV). Protein CDH2 (Cadherin 2), which plays a critical role in cell-to-cell junction formation^39^, was linked to both circulatory diseases (Chapter IX) and metabolic disorders (Chapter IV). Another interesting example is Pepsinogen A4 (PGA4), a key protein associated with metabolism^39^, which displayed prediction-specific associations with metabolic disorders (Chapter IV; Fig. 6D). Of note, PGA4 was also prioritized as the predictive biomarker for mental health disorders (Chapter V; Fig. 6D). Similar to the results derived from the diagnostic model, we also observed a biomarker synergy between Chapter IV and Chapter IX based on the predictive model (Fig. 6E). This prompted us to investigate the shared proteins and their functions: as expected, we recognized multiple proteins associated with vascular functions, including VWF, CDH2, and ANGPTL4^40^, which were detected by both models within the two disease categories (Fig. 6E).

**Fig. 6:**
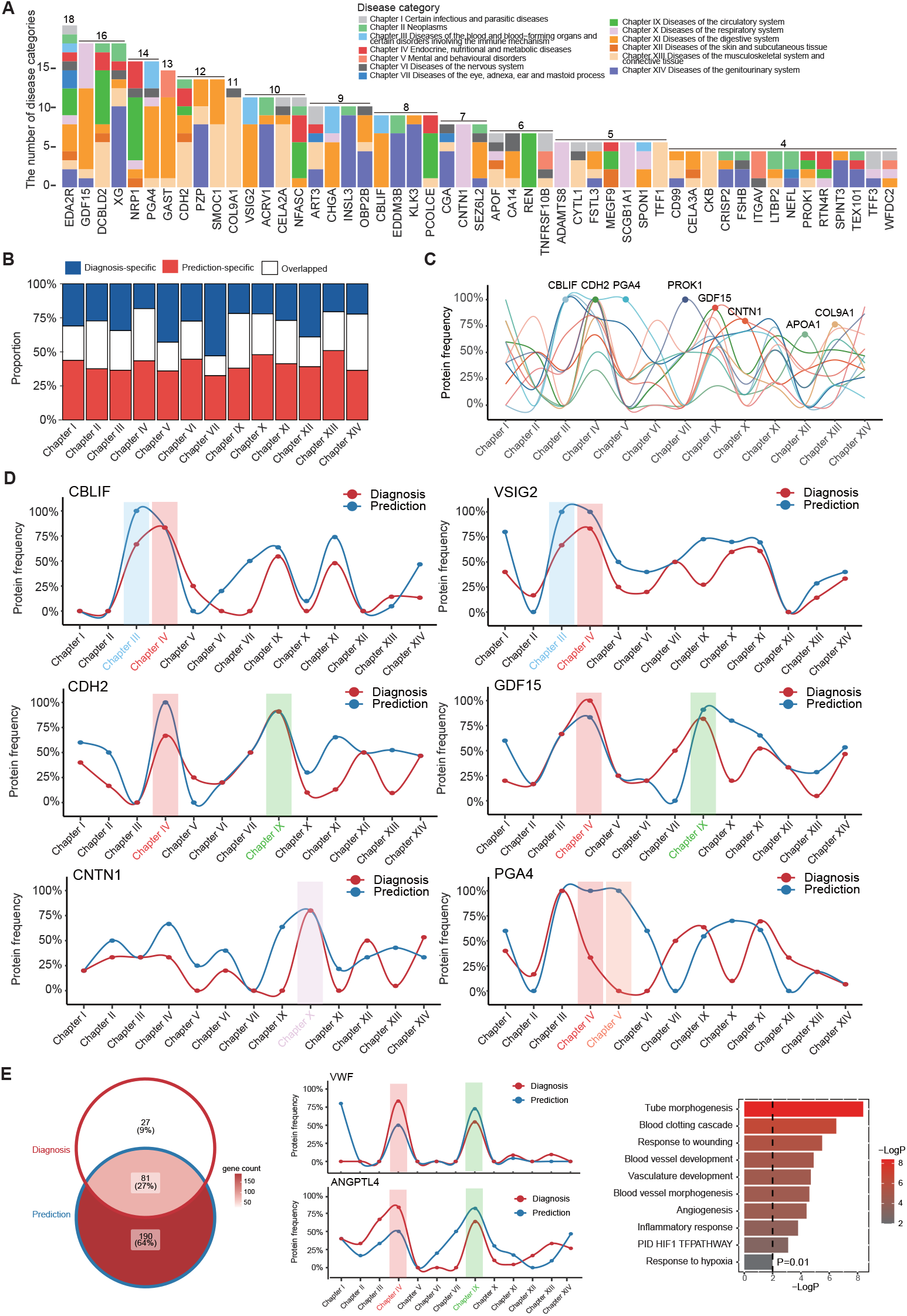
Prophet prioritizes plasma proteins related disease prediction. (A) Stacked bar chart of protein importance in predicting diseases, with colors representing different disease categories (throughout this figure). The number above each bar indicates the count of diseases in which the protein was ranked in the top 5. (B) Proportion of diagnosis/prediction-specific and overlapping proteins across various disease categories. (C) Line plot showing the frequencies of associated proteins in disease prediction across various disease categories. Representative proteins are highlighted with dots and labeled by their names. The frequency of a specific protein in a disease category was defined by the proportion of diseases within that category where the protein was identified as an associated factor. (D) Similar to (C), but for representative proteins showing differential associations between the diagnostic model and the predictive model. (E) Illustration and functional enrichment analysis of representative proteins shared between Chapter IV and Chapter IX. The Venn diagram illustrates the overlap of shared proteins between Chapters IV and IX, comparing the diagnostic model and the predictive model. *P*-value by hypergeometric test.

Taken together, these results highlight Prophet’s potential to discover biomarkers, informing disease mechanisms and new therapeutic targets. Where a single protein has relevance to multiple diseases, then there is significant translational potential, whether in the form of a therapeutic intervention, or a cost-efficient predictive biomarker.

### Prophet unveils proteomics-based disease similarities

Finally, we leveraged disease-specific query embeddings derived from Prophet to examine the proteomics-based disease similarities (Methods). Within expectation, we observed that diseases within the same categories exhibited stronger proteomic similarities than those across different groups for both models (Fig. 7A and 7B). In particular, the diagnostic Prophet model identified 59 disease pairs with a similarity greater than 0.9, and the predictive model identified 56 pairs above the same threshold (Supplementary Table 4). Of these, 27 pairs were shared by both models. These shared pairs typically showed either similar clinical definitions or strong mechanistic relationships. For example, disease pairs like “Meniscus derangement” and “Gonarthrosis” (diagnosis similarity = 0.908, prediction similarity = 0.935) and “Hyperplasia of prostate” and “Diseases of male genital organs” (diagnosis similarity = 0.958, prediction similarity = 0.968) presented strong diagnostic overlaps. Similarly, diseases such as “Menorrhagia” and “Polyp of the female genital tract” (diagnosis similarity = 0.938, prediction similarity = 0.956) and “Low back pain” and “Spondylopathies” (diagnosis similarity = 0.910, prediction similarity = 0.939) also showed close clinical symptoms. Interestingly, both of these relationships are potentially causal rather than correlative: polyps of the female genital tract are a cause of menorrhagia and spondyloarthropathy is a cause of back pain.

**Fig. 7:**
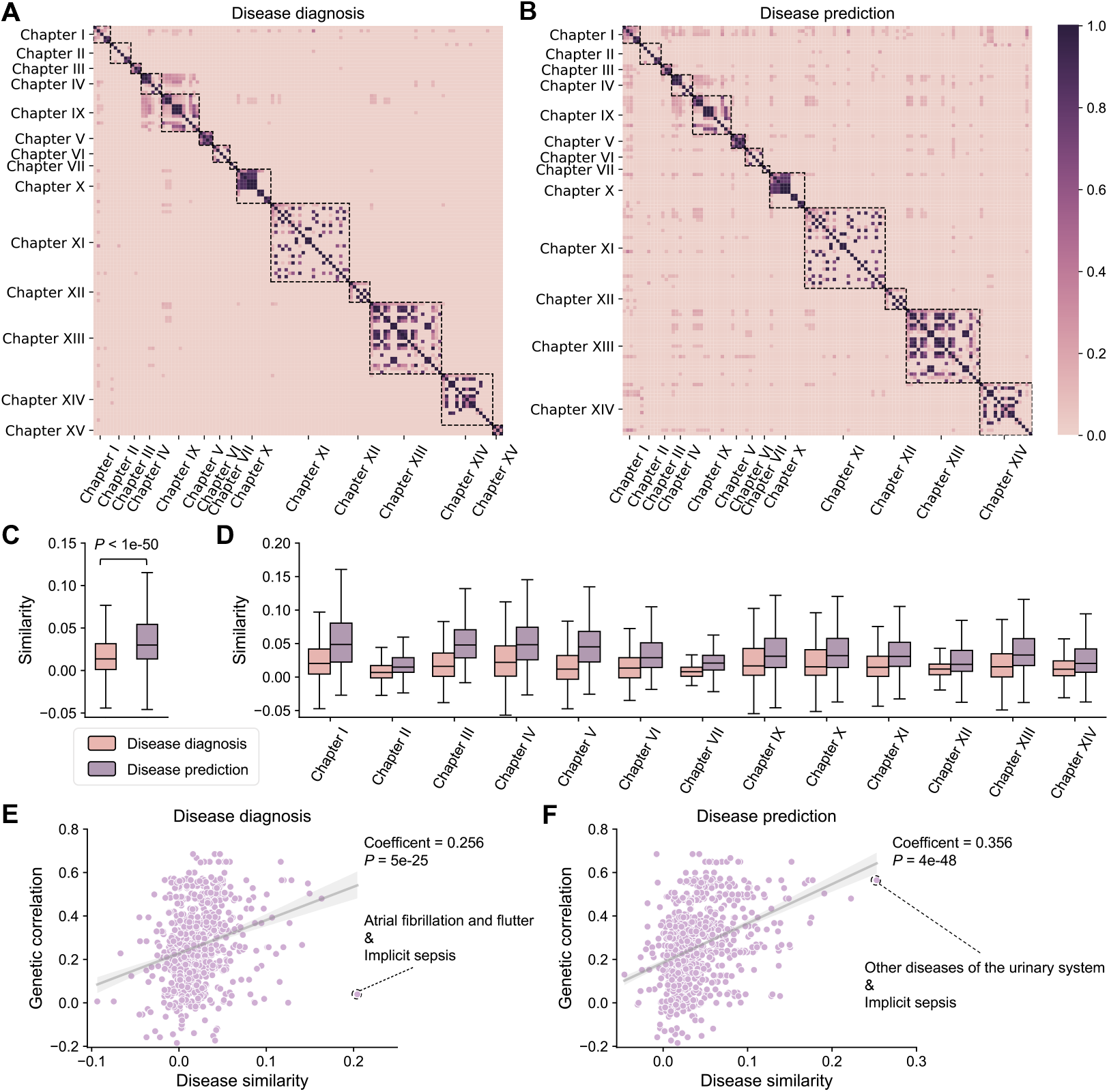
Prophet reveals proteomics-based disease similarities. (A-B) Visualizations of similarities between prevalent diseases (A) and incident diseases (B) learned by the Prophet model. The cosine similarity between disease query embeddings in the attentive readout module was used to measure the disease similarity. Disease groups were organized by their respective chapters and highlighted with dashed rectangles. (C) Comparison of average inter-group similarities between disease diagnostic and predictive models. *P*-value by two-sided paired *t*-test. (D) Inter-group similarity comparison between the diagnostic and predictive models across different disease categories. The box plot center line, limits, and whiskers represent the median, quartiles, and 1.5x interquartile range (IQR), respectively. (E-F) Correlation between genetic correlation and proteomic similarity of inter-group disease pairs for the diagnostic model (E) and the predictive model (F), respectively. Each dot represents a disease pair considered in the analysis. The genetic correlation *r*_*g*_ was estimated using the LD score regression (LDSC) based on the UKB data. The coefficient was computed by Pearson correlation. The linear regression line and a 95% confidence interval (CI) were also highlighted.

At the group level, the highest inter-group similarities were observed between Chapter IV (Endocrine, Nutritional and Metabolic Diseases) and Chapter IX (Diseases of the Circulatory System), with an average similarity of 0.138 (ranked #1) in the diagnostic model and 0.095 (ranked #2) with the predictive model. This suggested significant overlaps between diseases from these two categories. For instance, “Coronary atherosclerosis” and “Pure hypercholesterolaemia” had a similarity of 0.394 (ranked #2) in the diagnostic model and 0.201 (ranked #18) in the predictive model. Other pairs showed moderate similarities, such as “Metabolic disorders” and “Hypertension” with similarities of 0.312 (ranked #8) and 0.228 (ranked #12) in the diagnostic and predictive models, respectively.

Notably, we noticed that the disease similarities across different disease groups were significantly higher in the predictive model than in the diagnostic model (*P <* 1e-50, two-sided paired *t*-test; Fig. 7C and 7B). This trend was not observed for diseases within the same group (Supplementary Fig. 6). We wondered if the higher inter-group similarities with the predictive model might be a result of shared underlying causal biological factors, rather than a reflection of shared symptoms. To investigate this, we compared proteomic similarity scores to genetic correlations between diseases^41^ (Methods), wherein the latter is commonly used to tag shared upstream disease underpinnings^42^. As predicted, the correlation between predictive-model-derived proteomic similarities and genetic correlations was markedly higher than for diagnostic-model-based proteomic similarities (Pearson correlation coefficient 0.356 versus 0.256; Fig. 7E and 7F).

The highest inter-group diagnostic similarity was between “Atrial fibrillation and flutter” and “Implicit sepsis” (similarity = 0.2; Supplementary Table 4). Atrial fibrillation/flutter is a common symptom of sepsis, but most patients with fibrillation/flutter do not have sepsis; this is reflected in a low genetic correlation of only 0.04. In contrast, the largest inter-group predictive similarity was between “Other diseases of the urinary system” and “Implicit sepsis” (similarity = 0.25; Supplementary Table 4), reinforced by their strong genetic correlation of 0.56. This indicates a causative relationship between abnormalities of the urinary tract and urinary sepsis which is sufficiently common to necessitate clinical action^43^.

## Discussion

In this study, we present Prophet, a deep learning-based framework designed to enhance both disease diagnosis and prediction while offering versatile interpretability. By utilizing a multistage, multitask training strategy, Prophet effectively captures the complex relationships between the proteome and the phenome. The incorporation of the attentive readout module further enhances model interpretability, enabling simultaneous disease prediction and biological discovery.

Through extensive testing on 120 prevalent and 117 incident diseases, Prophet was demonstrated to consistently outperform all baseline methods, improving disgnostic and predictive performance for almost all diseases. Additionally, we validated Prophet’s effectiveness in disease risk stratification, reinforcing its potential as a clinical tool to advance precision medicine. We showed that disease predictions made with Prophet have the potential to advance translational medicine by uncovering new broadly-relevant proteomic biomarkers, therapeutic targets, and causal relationships between diseases.

Clinical covariates such as age, sex, medical history, and other features from EHRs have been widely used for disease diagnosis and prediction^44^. These clinical factors are essential as they provide valuable indicators for disease risk or phenotypes for individuals. Age and sex are known to influence disease likelihood, and comorbidities or medication usage can affect disease progression. Although Prophet is a proteomics-based model, it can be extended to incorporate these clinical features without difficulties. The integration of multi-modality data would allow the model to yield more personalized and accurate predictions, potentially improving its clinical relevance and utility, especially when updated in real-time through EHRs. On the other hand, clinical features could also be correlated with proteomic profiles. Indeed, we showed that Prophet yielded sex-specific predictions for certain diseases, suggesting that our model captured sex-dependent proteomic signatures.

Biomarkers identified by Prophet represent proteome-wide signatures of diseases, either upstream or downstream of disease pathogenesis. Although in our study, we linked Prophet discoveries, especially those derived from the predictive model, to genetic signals, the causal factors could not be precisely pinpointed without supports from additional functional data. Specifically, genomic, transcriptomic, and epigenomic data provide complementary insights into biological mechanisms which demographic and clinical data cannot offer. For example, protein quantitative trait loci^45^ (pQTLs) analysis associates genetic variants to the variation of protein expression which further impact individuals’ phenotypes. This linkage better maps the influence of proteomic changes on phenotypes than solely based on proteomic profiling. Therefore, to interpret Prophet’s biomarkers towards mechanisms, we recommend integrating other types of functional data to control the false positive rate.

In summary, Prophet represents a potent and versatile framework for individualized proteomic data analysis, enabling precise disease diagnosis, disease prediction, and systematic biomarker discovery. Our work provides a methodological foundation for proteomics-based precision medicine.

## Methods

### Data collection and preprocessing

#### Plasma proteomics dataset

The data analyzed in this study were obtained from UKB^46^, a largescale, population-based biobank that includes approximately 500,000 participants aged from 40 to 69 years, recruited between 2006 and 2010 across 22 recruitment centers in the UK. We utilized proteomics data from UKB-PPP^8^, which measured 2,924 unique proteins in plasma samples from 53,014 UKB participants. The proteomic profiling was conducted using the Olink Explore Proximity Extension Assay with next-generation sequencing. Plasma samples were processed, stored at − 80°C, and subsequently analyzed for proteins related to cardiometabolic, inflammation, neurology, and oncology pathways. Rigorous quality control measures were employed, including the normalization of protein expression through normalized protein expression (NPX) values to minimize batch effects and improve accuracy, particularly for low-abundance proteins. For further details on sample selection, processing, and quality control, please refer to the relevant publications^8, 47^.

#### Disease definition

The UKB diagnosis data were extracted from the hospital inpatient records (Fields 41270 and 41280 in Category 2000), classified using the International Classification of Diseases (ICD)-10 codes. Following the state-of-the-art practice^17^, we adopted the endpoint and control definitions in FinnGen Data Freeze 12, and followed the FinnGen’s quality control guidelines^24^. Diseases were categorized as either prevalent or incident, based on whether they occurred before or after participants’ baseline visits, at which time blood samples were collected. In the analysis of incident diseases, participants diagnosed with the disease before the collection of blood samples were excluded. Similarly, for prevalent diseases, participants diagnosed during follow-ups were excluded. Controls for each condition were defined as participants who did not have that disease at any point during the study period. To ensure sufficient sample size and robust training and evaluation, we excluded endpoints with fewer than 500 cases for prevalent diseases and fewer than 200 cases for incident diseases. The final analysis-ready dataset included 120 prevalent and 117 incident disease endpoints for the study cohort with available proteomics data (Supplementary Tables 1 and 2). In addition, participants with missing data in Field 41280 were excluded, resulting in a final cohort of 53,014 participants. The normalized expression levels for 2,924 proteins were used as the model input.

### Prophet model details

In this section, we present Prophet (Fig. 1), which consists of the proteomics transformer architecture, a self-supervised pretraining strategy, and a prompt-based continuous fine-tuning approach for prevalent and incident disease prediction.

### Proteomics transformer

#### Transformer encoder

The transformer encoder processes the proteomics data vector **X** ∈ ℝ^*N*×1^, where each element represents the measurement of the *i*-th protein and *N* represents the total number of proteins, and outputs corresponding protein embeddings. The raw protein measurements **X** are first projected into a high-dimensional space through an embedding layer, computed as

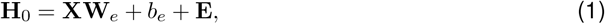

where **W**_*e*_ is the embedding weight matrix of size 1 *× d*_model_ and *b*_*e*_ is the bias term of size *d*_model_. 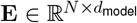 is a trainable matrix representing the protein-specific positional embeddings, which enables the model to retain the distinct identity of each protein. The resulting matrix 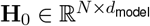 is the initial protein embeddings and serves as the input to subsequent layers of the transformer encoder.

Each transformer encoder layer consists of two main components: (1) a multi-head self-attention mechanism and (2) a position-wise feed-forward network. In each self-attention layer, the input embeddings **H**_*l*_ (where *l* is the layer index) are transformed into three distinct matrices, i.e., the query **Q** = **H**_*l*_**W**_*Q*_, the key **K** = **H**_*l*_**W**_*K*_, and the value **V** = **H**_*l*_**W**_*V*_, with **W**_*Q*_, **W**_*K*_, and **W**_*V*_ being trainable weight matrices of size *d*_model_ × *d*_model_. The attention scores are then computed as

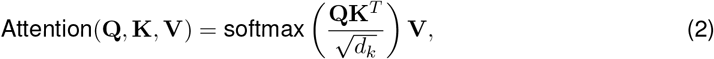

where *d*_*k*_ is the dimension of the query and key vectors.

After the self-attention step, the output is passed through a position-wise feed-forward network (FFN) consisting of two fully connected layers with a ReLU activation function^48^:

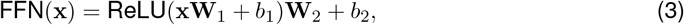

where **W**_1_ and **W**_2_ are weight matrices of size *d*_model_ *× d*_ffn_ and *d*_ffn_ *× d*_model_, respectively, and *b*_1_ and *b*_2_ are bias terms. The output of this feed-forward network is then combined with the input via a residual connection, followed by layer normalization^49^.

This process is iteratively conducted across multiple layers of the transformer encoder, with each layer progressively refining the protein representations. As the data flows through successive transformer layers, the model is able to capture increasingly complex and non-linear interactions among proteins. The final output, denoted as **H**_*L*_, where *L* indicates the total number of encoder layers, is then used for downstream tasks such as prevalent and incident disease prediction.

#### Attentive readout module

After the transformer encoder layers, the encoded protein embeddings **H**_*L*_ are passed to an attentive readout module, which summarizes the protein representations by focusing on the proteins that contribute most to the prediction. We implement an attention mechanism that dynamically adjusts the importance of each protein for different diseases.

In particular, each disease has a trainable query embedding 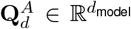, which is used to compute disease-specific attention scores for proteins. The attention module computes a weighted sum of protein embeddings, with the weight assigned to each protein reflecting its relevance to a specific disease. Initially, the protein embeddings **H**_*L*_ are transformed into a key matrix, denoted as **K**^*A*^, and a value matrix, denoted as **V**^*A*^. It is important to note that both **K**^*A*^ and **V**^*A*^ are shared across all diseases.

The attention scores **A**_*d*_ for each protein-disease pair are computed by the dot product of the keys **K**^*A*^ and the query vectors 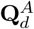 with a cosine similarity function:

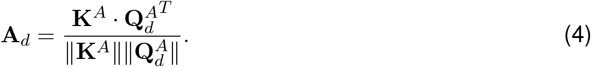

The attention scores **A**_*d*_ ∈ ℝ^*N*^ estimate the importance of each protein in relation to different diseases.

The weighted sum of the value embeddings **V** based on the disease-specific attention scores is calculated as:

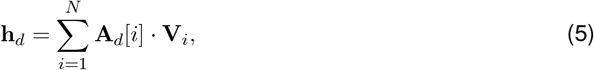

where **h**_*d*_ ∈ ℝ^1*×D*^ stands for the pooled protein embeddings for disease *d*.

Finally, the disease-specific embeddings **h**_*d*_ are used to predict disease phenotypes via a disease-specific linear layer:

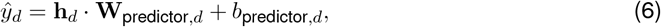

where **W**_predictor,*d*_ and *b*_predictor,*d*_ are trainable weight and bias terms, respectively, and *ý*_*d*_ is the predicted disease risk score.

The attention module allows for the prioritization of important proteins for disease prediction, thereby improving model interpretability and the identification of key biomarkers. Further, by utilizing disease-specific query embeddings, Prophet supports multitask learning and enables the sharing of common knowledge across tasks, ultimately enhancing overall performance.

### Model training

#### Stage 1: Self-supervised pretraining

During pretraining, a random subset of non-zero protein measurements is masked. The model is then tasked with predicting these masked values using the context of unmasked proteins. To achieve this, the transformer encoder processes the input data to generate protein embeddings, which are subsequently transformed by a multi-layer perceptron (MLP) to predict the expression values of masked proteins. We use the mean squared error (MSE) between the predicted and measured protein expression values as the loss function.

#### Stage 2: Prompt-based fine-tuning for prevalent disease prediction

In this approach, trainable prompt embeddings are introduced alongside the initial protein embeddings, which are used together as input to the transformer encoder.

Specifically, a set of trainable prompt vectors 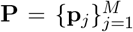 is defined, where *M* denotes the number of prompt vectors and each prompt vector **p**_*j*_ has a dimension of *d*_model_. These prompts are concatenated with the initial protein embeddings **H**_0_, resulting in an augmented input matrix:

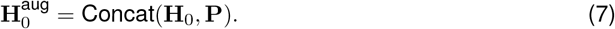

This augmented matrix is next fed into the pretrained transformer encoder. At each layer *l* of the encoder, the input is updated as follows:

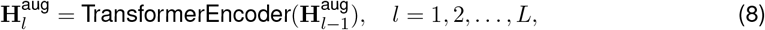

where 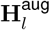 represents the embeddings after the *l*-th layer, and *L* is the total number of layers in the transformer encoder. The final embeddings 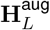 are fed into an attentive readout module to make predictions for all prevalent diseases in a multitask learning setting. The binary cross-entropy (BCE) loss is used to optimize the model parameters, defined as:

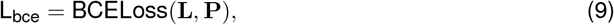

where **L** and **P** represent the label and prediction vectors, respectively.

#### Disease cooccurrence regularization

Additionally, we propose a disease cooccurrence regularization loss that penalizes discrepancies between the similarity of disease-specific query embeddings 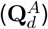 and the disease cooccurrence. The primary objective is to ensure that similar diseases are consistently represented in the embedding space. Specifically, for each sample, we first calculate the cooccurrence matrix **C**, which captures the pairwise similarity between disease labels **L**.

The cooccurrence matrix is defined as:

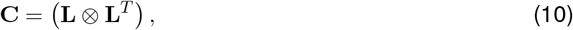

where ⊗ denotes the element-wise equality operator. This results in a binary matrix where **C**_*i*,*j*_ = 1 if diseases *i* and *j* co-occur for this sample, and **C**_*i*,*j*_ = 0 otherwise. The disease similarity matrix is then defined as:

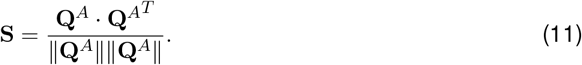

To focus on meaningful disease pairs, we mask both the cooccurrence matrix **C** and the similarity matrix **S** to ignore irrelevant label pairs. In particular, we define the binary mask **M** as:

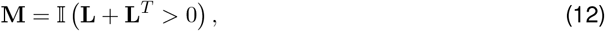

where 𝕀 is the indicator function. This masking matrix ensures that only pairs where at least one disease is present are considered in the loss. The masked matrices are then given by:

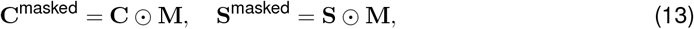

where ⊙ denotes element-wise multiplication.

The disease cooccurrence penalty is computed as the mean squared difference between the masked similarity matrix **S**^masked^ and the masked cooccurrence matrix **C**^masked^. For each sample, the penalty is given by:

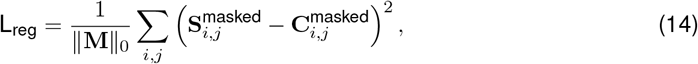

where ∥**M**∥_0_ is the number of valid label pairs defined by the masking matrix.

The final loss function per sample is given by:

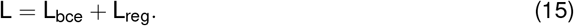

We note that, during stage 2, the pretrained transformer encoder remains fixed, but only the parameters of the prompt embeddings and the attentive readout module are optimized using this loss. This approach helps the model retain as much knowledge as possible learned from pretraining.

#### Stage 3: Prompt-based fine-tuning for incident disease prediction

Building on the model finetuned for prevalent disease prediction, we further fine-tune Prophet for incident disease prediction. In this process, all parameters from stage 2 are carried over to stage 3. The transformer encoder remains fixed, while only the prompt embeddings and the attentive readout module are optimized using the same loss function as in stage 2.

Importantly, to prevent information leakage, the same training, validation, and testing dataset split was used throughout all three stages of model training.

### Model evaluation and benchmarking

In this study, we used UKB-PPP proteomics data, comprising 53,014 participants and measurements of 2,924 proteins, to predict 120 prevalent and 117 incident diseases (see Data collection and preprocessing). The dataset was randomly partitioned it into training, validation, and test sets with a ratio of 8:1:1. The validation dataset was used for early stopping and the final performance was evaluated on the test dataset.

To benchmark the performance of our proposed framework, we also implemented six baseline methods, including both linear and non-linear models. The linear models included linear regression and Lasso, and the non-linear models encompassed K-nearest neighbors (KNN), random forest, multilayler perceptron (MLP), and XGBoost^32^. The key hyperparameters of these baseline methods were optimized based on the held-out validation dataset.

We adopted two metrics – the area under the receiver operating characteristic curve (AUROC) and the area under precision-recall curve (AUPRC) – to measure the performance of binary classification. Predictive performance of all methods was estimated based on five independent repeats with different random seeds for both data partitioning and model initialization. The same training, validation, and testing dataset splits were used across Prophet and all baseline models for a fair comparison.

### Implementation details

Prophet was mainly implemented in PyTorch 1.13.1 with CUDA 11.6 and Python 3.7.16. We implemented a six-layer transformer encoder with a hidden size of 128 and two attention heads. We set the number of prompts to six and the disease cooccurrence regulation coefficient to one. An AdamW optimizer with a learning rate of 4e-4 was used to train model. The training was performed with a batch size of 128 for a total of 50 epochs, employing early stopping. The experiments were carried out using two NVIDIA A100 GPUs.

Baseline models were implemented based on Scikit-learn 1.0.2 and XGBoost (https://xgboost.ai/).

### Risk stratification analysis

We defined high-risk individuals as those individuals whose prediction scores were in the top 5% of all predictions and low-risk individuals as those in the bottom 5%. Additionally, we considered the overall incidence rate of the disease in the UKB dataset as the background for comparison.

To quantitatively evaluate the performance of Prophet for risk stratification, we used the area under the cumulative incidence curve (AUCIC). Specifically, for high-risk or low-risk group, we performed Kaplan-Meier (KM) survival analysis to estimate the cumulative incidence of the disease. The Kaplan-Meier estimator provides a non-parametric way to estimate the cumulative probability of disease occurrence over time, producing a cumulative incidence curve that reflects the probability of disease occurrence at each time point. Next, we calculated the AUCIC by summing up the areas of trapezoids formed between adjacent points along the cumulative incidence curve. To ensure comparability across models, the AUCIC values were normalized by dividing the computed AUCIC by the maximum possible area under the curve, which corresponded to a model that predicted the disease occurrence at every time point with certainty. This normalization resulted in AUCIC values ranging from 0 to 1.

### Identification of disease-associated proteins based on Prophet

To identify protein-disease associations, we used a two-step statistical approach. First, for each disease, we calculated *Z*-scores for protein attention scores in both case and control groups, normalized by their mean and standard deviation. We then computed two-tailed *P*-values based on these *Z*-scores to assess differential attention scores, in which a threshold of 0.05 was applied to select significant proteins for each individual. Next, we carried out two-sided Fisher’s exact test for each protein to determine its association with the disease. To account for multiple comparisons, we applied the Benjamini-Hochberg (BH) procedure to control the false discovery rate (FDR), and used an FDR *<* 0.01 to derive the final significant protein-disease associations.

### Sex difference analysis

To examine the impact of sex on Prophet’s prediction, we first calculated the incidence rates across every 5th percentile of Prophet’s predictions for both males and females. Next, we performed a two-sided Wilcoxon signed-rank test to determine whether there was a significant difference between males and females. The BH procedure with an FDR threshold of *<*0.05 was applied to control the FDR in multiple comparisons.

#### Gene set functional enrichment analysis

Functional enrichment analysis for protein sets of interest was conducted using the Metascape^50^ webtool (https://metascape.org/). Function terms with a *P*-value*<*0.01 were considered to be significantly enriched. Representative pathways and/or gene sets were selected for illustration.

#### Genetic correlation analysis

Genetic correlation scores were obtained from https://ukbb-rg.hail.is/, in which the disease genetic correlation *r*_*g*_ was computed using the LD score regression^41^ (LDSC) based on the UKB data. Only overlapping disease pairs between our data and the genetic correlation data were included in the analysis.

## Data availability

All proteomic, phenotypic, and EHR data used in this study are available from UKB upon application (https://www.ukbiobank.ac.uk).

## Code availability

The source code and tutorial of Prophet are available at GitHub (https://github.com/lihan97/Prophet).

## Acknowledgments

We thank the UKB participants for sharing their data, which were accessed under the application no. 101835. This work was supported by the National Key R&D Program of China (2024YFC3407800 to H.L. and X.X.) and the Strategic Priority Research Program of Chinese Academy of Sciences (XDA0460203 to W.Z.).

## Author contributions

H.L. and S.Z. conceived the concept and designed the study. H.L. and Y.Li developed Prophet and performed data analysis. H.L., Y.Li, Y.Liu, J.C.-K., P.G., X.S., S.C., X.X. and S.Z. are responsible for data interpretation. S.Z., X.X., S.C. and H.L. supervised the project. H.L. and S.Z. prepared the manuscript with assistance from all other authors.

## Supplementary Figures

**Supplementary Fig. 1**: Performance evaluation for disease diagnosis in AUPRC. (A) AUPRC scores of Prophet and six baseline methods for predicting 120 prevalent diseases. *P*-value by paired *t*-test. The bar plot and error bar denote the mean and standard deviation, respectively. AUPRC, area under the precision-recall curve; MLP, multilayer perceptron; KNN, *k*-nearest neighbors. (B) AUPRC comparison between Prophet (red) and XGBoost (blue) across different disease categories. The training and testing procedure was conducted for five repeats using different random seeds for all models.

**Supplementary Fig. 2**: Ablation study of Prophet. (A) Comparison of AUROC (top) and AUPRC (bottom) between Prophet and its variants, including Prophet without prompt-based fine-tuning, Prophet without disease cooccurrence regularization, and Prophet without pretraining. The bar plot and error bar denote the mean and standard deviation, respectively. AUROC, area under the receiver operating characteristic curve; AUPRC, area under the precision-recall curve. (B) Comparison of AUROC (top) and AUPRC (bottom) for Prophet with varying numbers of prompts. The dot and shaded area represent the mean and standard deviation, respectively. (C) Comparison of AUROC (top) and AUPRC (bottom) for Prophet with different disease cooccurrence regularization coefficients. The training and testing procedure was conducted for five repeats using different random seeds for all models.

**Supplementary Fig. 3**: Performance evaluation for disease prediction in AUPRC. (A) AUPRC scores of Prophet and six baseline methods for predicting 117 incident diseases. *P*-value by paired *t*-test. The bar plot and error bar denote the mean and standard deviation, respectively. AUPRC, area under the precision-recall curve; MLP, multilayer perceptron; KNN, *k*-nearest neighbors. (B) AUPRC comparison between Prophet (red) and XGBoost (blue) across different disease categories. The training and testing procedure was conducted for five repeats using different random seeds for all models.

**Supplementary Fig. 4**: Evaluation of risk stratification based on Prophet for disease diagnosis. (A) Model performance for disease diagnosis across different time windows, which were calculated based on the time difference between diagnosis and the baseline visit. The positive-to-negative ratio was ensured to be consistent across time intervals. The dot and the shaded region denote the mean and standard deviation, respectively. AUROC, area under the receiver operating characteristic curve; AUPRC, area under the precision-recall curve. (B) Comparison of the area under the cumulative incidence curve (AUCIC) between Prophet and XGBoost for high-risk individuals (right) and low-risk individuals (left). High-risk and low-risk individuals are defined as those individuals whose risk scores of prevalent diseases are in the top 5% and bottom 5% of all predictions, respectively. Each dot represents a disease. (C) Incidence rates for the eight diseases with the largest difference between sexes, against Prophet’s prediction percentiles based on the diagnostic model. *P*-value by two-sided Wilcoxon signed-rank test. The dot and shaded area represent the mean and standard deviation, respectively. The male background (green dashed line) and female background (red dashed line) denote the estimated incidence rates of males and females, respectively.

**Supplementary Fig. 5**: The cumulative incidence curves for high-risk and low-risk individuals of G6 NERPLEX (Nerve, nerve root and plexus disorders), as identified by the (A) disease diagnostic models and (B) disease predictive models. The background curve (gray) denotes the incidence rates estimated based on the whole dataset over time.

**Supplementary Fig. 6**: Intra-group disease similarity analysis. (A) Comparison of average intragroup disease similarities between the disease diagnostic and predictive models. (B) Intra-group similarity comparison between the diagnostic and predictive models across different disease categories. The box plot center line, limits, and whiskers represent the median, quartiles, and 1.5x interquartile range (IQR), respectively.

## Supplementary Tables

**Supplementary Table 1**: List of prevalent and incident diseases included in this study, based on the definitions from FinnGen Data Freeze 12.

**Supplementary Table 2**: AUROC, AUPRC, and AUCIC scores of Prophet for disease diagnosis and prediction. AUROC, area under the receiver operating characteristic curve; AUPRC, area under the precision-recall curve; AUCIC, area under the accumulative incidence curve.

**Supplementary Table 3**: Protein-disease associations identified by Prophet in disease diagnosis and prediction.

**Supplementary Table 4**: Proteomics-based disease similarities calculated based on the disease diagnostic and predictive models.

## References

1. Richard Hodson. Precision medicine. Nature, 537(7619):S49–S49, 2016.

2. Euan A Ashley. Towards precision medicine. Nature Reviews Genetics, 17(9):507–522, 2016.

3. Joshua C Denny and Francis S Collins. Precision medicine in 2030—seven ways to transform healthcare. Cell, 184(6):1415–1419, 2021.

4. Elena Fountzilas, Apostolia M Tsimberidou, Henry Hiep Vo, and Razelle Kurzrock. Clinical trial design in the era of precision medicine. Genome medicine, 14(1):101, 2022.

5. Ashley J Vargas and Curtis C Harris. Biomarker development in the precision medicine era: lung cancer as a case study. Nature reviews cancer, 16(8):525–537, 2016.

6. Dearbhaile C Collins, Raghav Sundar, Joline SJ Lim, and Timothy A Yap. Towards precision medicine in the clinic: from biomarker discovery to novel therapeutics. Trends in pharmacological sciences, 38(1):25–40, 2017.

7. Wayne C Drevets, Gayle M Wittenberg, Edward T Bullmore, and Husseini K Manji. Immune targets for therapeutic development in depression: towards precision medicine. Nature reviews Drug discovery, 21(3):224–244, 2022.

8. Benjamin B Sun, Joshua Chiou, Matthew Traylor, Christian Benner, Yi-Hsiang Hsu, Tom G Richardson, Praveen Surendran, Anubha Mahajan, Chloe Robins, Steven G Vasquez-Grinnell, et al. Plasma proteomic associations with genetics and health in the uk biobank. Nature, 622(7982):329–338, 2023.

9. Philipp E Geyer, Nils A Kulak, Garwin Pichler, Lesca M Holdt, Daniel Teupser, and Matthias Mann. Plasma proteome profiling to assess human health and disease. Cell systems, 2(3):185–195, 2016.

10. Stephen A Williams, Mika Kivimaki, Claudia Langenberg, Aroon D Hingorani, Juan P Casas, Claude Bouchard, Christian Jonasson, Mark A Sarzynski, Martin J Shipley, Leigh Alexander, et al. Plasma protein patterns as comprehensive indicators of health. Nature medicine, 25(12):1851–1857, 2019.

11. Wen Zhong, Fredrik Edfors, Anders Gummesson, Göran Bergström, Linn Fagerberg, and Mathias Uhlén. Next generation plasma proteome profiling to monitor health and disease. Nature communications, 12(1):2493, 2021.

12. Ryan S Dhindsa, Oliver S Burren, Benjamin B Sun, Bram P Prins, Dorota Matelska, Eleanor Wheeler, Jonathan Mitchell, Erin Oerton, Ventzislava A Hristova, Katherine R Smith, et al. Rare variant associations with plasma protein levels in the uk biobank. Nature, 622(7982):339–347, 2023.

13. Jia You, Yu Guo, Yi Zhang, Ju-Jiao Kang, Lin-Bo Wang, Jian-Feng Feng, Wei Cheng, and Jin-Tai Yu. Plasma proteomic profiles predict individual future health risk. Nature Communications, 14(1):7817, 2023.

14. Julia Carrasco-Zanini, Maik Pietzner, Jonathan Davitte, Praveen Surendran, Damien C Croteau-Chonka, Chloe Robins, Ana Torralbo, Christopher Tomlinson, Florian Grünschläger, Natalie Fitzpatrick, et al. Proteomic signatures improve risk prediction for common and rare diseases. Nature medicine, 30(9):2489–2498, 2024.

15. M Austin Argentieri, Sihao Xiao, Derrick Bennett, Laura Winchester, Alejo J Nevado-Holgado, Upamanyu Ghose, Ashwag Albukhari, Pang Yao, Mohsen Mazidi, Jun Lv, et al. Proteomic aging clock predicts mortality and risk of common age-related diseases in diverse populations. Nature medicine, 30(9):2450–2460, 2024.

16. Yu Guo, Jia You, Yi Zhang, Wei-Shi Liu, Yu-Yuan Huang, Ya-Ru Zhang, Wei Zhang, Qiang Dong, Jian-Feng Feng, Wei Cheng, et al. Plasma proteomic profiles predict future dementia in healthy adults. Nature Aging, 4(2):247–260, 2024.

17. Yue-Ting Deng, Jia You, Yu He, Yi Zhang, Hai-Yun Li, Xin-Rui Wu, Ji-Yun Cheng, Yu Guo, Zi-Wen Long, Yi-Lin Chen, et al. Atlas of the plasma proteome in health and disease in 53,026 adults. Cell, 2024.

18. Art Schuermans, Ashley B. Pournamdari, Jiwoo Lee, Rohan Bhukar, Shriienidhie Ganesh, Nicholas Darosa, Aeron M. Small, Zhi Yu, Whitney Hornsby, Satoshi Koyama, Charles Kooperberg, Alexander P. Reiner, James L. Januzzi, Michael C. Honigberg, and Pradeep Natarajan. Integrative proteomic analyses across common cardiac diseases yield mechanistic insights and enhanced prediction. Nature Cardiovascular Research, 3(12):1516–1530, Dec 2024.

19. Heather J Cordell. Detecting gene–gene interactions that underlie human diseases. Nature Reviews Genetics, 10(6):392–404, 2009.

20. Haiying Lu, Qiaodan Zhou, Jun He, Zhongliang Jiang, Cheng Peng, Rongsheng Tong, and Jianyou Shi. Recent advances in the development of protein–protein interactions modulators: mechanisms and clinical trials. Signal transduction and targeted therapy, 5(1):213, 2020.

21. Damian Szklarczyk, Rebecca Kirsch, Mikaela Koutrouli, Katerina Nastou, Farrokh Mehryary, Radja Hachilif, Annika L Gable, Tao Fang, Nadezhda T Doncheva, Sampo Pyysalo, et al. The string database in 2023: protein–protein association networks and functional enrichment analyses for any sequenced genome of interest. Nucleic acids research, 51(D1):D638–D646, 2023.

22. Jack F Greenblatt, Bruce M Alberts, and Nevan J Krogan. Discovery and significance of protein-protein interactions in health and disease. Cell, 187(23):6501–6517, 2024.

23. A Vaswani. Attention is all you need. Advances in Neural Information Processing Systems, 2017.

24. Mitja I Kurki, Juha Karjalainen, Priit Palta, Timo P Sipilä, Kati Kristiansson, Kati M Donner, Mary P Reeve, Hannele Laivuori, Mervi Aavikko, Mari A Kaunisto, et al. Finngen provides genetic insights from a well-phenotyped isolated population. Nature, 613(7944):508–518, 2023.

25. Minsheng Hao, Jing Gong, Xin Zeng, Chiming Liu, Yucheng Guo, Xingyi Cheng, Taifeng Wang, Jianzhu Ma, Xuegong Zhang, and Le Song. Large-scale foundation model on single-cell transcriptomics. Nature Methods, pages 1–11, 2024.

26. Hugo Dalla-Torre, Liam Gonzalez, Javier Mendoza-Revilla, Nicolas Lopez Carranza, Adam Henryk Grzywaczewski, Francesco Oteri, Christian Dallago, Evan Trop, Bernardo P de Almeida, Hassan Sirelkhatim, et al. Nucleotide transformer: building and evaluating robust foundation models for human genomics. Nature Methods, pages 1–11, 2024.

27. Xi Fu, Shentong Mo, Alejandro Buendia, Anouchka P Laurent, Anqi Shao, Maria del Mar Alvarez-Torres, Tianji Yu, Jimin Tan, Jiayu Su, Romella Sagatelian, et al. A foundation model of transcription across human cell types. Nature, pages 1–9, 2025.

28. Yanrong Ji, Zhihan Zhou, Han Liu, and Ramana V Davuluri. Dnabert: pre-trained bidirectional encoder representations from transformers model for dna-language in genome. Bioinformatics, 37(15):2112–2120, 2021.

29. Fan Yang, Wenchuan Wang, Fang Wang, Yuan Fang, Duyu Tang, Junzhou Huang, Hui Lu, and Jianhua Yao. scbert as a large-scale pretrained deep language model for cell type annotation of single-cell rna-seq data. Nature Machine Intelligence, 4(10):852–866, 2022.

30. Han Li, Ruotian Zhang, Yaosen Min, Dacheng Ma, Dan Zhao, and Jianyang Zeng. A knowledge-guided pre-training framework for improving molecular representation learning. Nature Communications, 14(1):7568, 2023.

31. Pengfei Liu, Weizhe Yuan, Jinlan Fu, Zhengbao Jiang, Hiroaki Hayashi, and Graham Neubig. Pre-train, prompt, and predict: A systematic survey of prompting methods in natural language processing. ACM Computing Surveys, 55(9):1–35, 2023.

32. Tianqi Chen and Carlos Guestrin. Xgboost: A scalable tree boosting system. In Proceedings of the 22nd acm sigkdd international conference on knowledge discovery and data mining, pages 785–794, 2016.

33. Georgios Voloudakis, James M Vicari, Sanan Venkatesh, Gabriel E Hoffman, Kristina Dobrindt, Wen Zhang, Noam D Beckmann, Christina A Higgins, Stathis Argyriou, Shan Jiang, et al. A translational genomics approach identifies il10rb as the top candidate gene target for covid-19 susceptibility. NPJ Genomic Medicine, 7(1):52, 2022.

34. Andrea Mattiotti, Stuti Prakash, Phil Barnett, and Maurice JB van den Hoff. Follistatin-like 1 in development and human diseases. Cellular and Molecular Life Sciences, 75:2339–2354, 2018.

35. John R Petrie, Tomasz J Guzik, and Rhian M Touyz. Diabetes, hypertension, and cardiovascular disease: clinical insights and vascular mechanisms. Canadian Journal of Cardiology, 34(5):575–584, 2018.

36. Guanghong Jia and James R Sowers. Hypertension in diabetes: an update of basic mechanisms and clinical disease. Hypertension, 78(5):1197–1205, 2021.

37. Connie B Newman, Michael J Blaha, Jeffrey B Boord, Bertrand Cariou, Alan Chait, Henry G Fein, Henry N Ginsberg, Ira J Goldberg, M Hassan Murad, Savitha Subramanian, and Lisa R Tannock. Lipid management in patients with endocrine disorders: An endocrine society clinical practice guideline. The Journal of Clinical Endocrinology Metabolism, 105(12):3613–3682, 09 2020.

38. Patryk Mucha, Filip Kus, Dominik Cysewski, Ryszard T Smolenski, and Marta Tomczyk. Vitamin b12 metabolism: a network of multi-protein mediated processes. International Journal of Molecular Sciences, 25(15):8021, 2024.

39. Xiaoling Gao, Yanjuan Jia, Hui Xu, Yonghong Li, Qing Zhu, Chaojun Wei, Jinxia Hou, Dehong Li, Wanxia Wang, Zhenhao Li, et al. Association between serum pepsinogen and atherosclerotic cardiovascular disease. Nutrition, Metabolism and Cardiovascular Diseases, 31(1):169– 177, 2021.

40. Felice Gragnano, Simona Sperlongano, Enrica Golia, Francesco Natale, Renatomaria Bianchi, Mario Crisci, Fabio Fimiani, Ivana Pariggiano, Vincenzo Diana, Andreina Carbone, et al. The role of von willebrand factor in vascular inflammation: from pathogenesis to targeted therapy. Mediators of inflammation, 2017(1):5620314, 2017.

41. Brendan K. Bulik-Sullivan, Po-Ru Loh, Hilary K. Finucane, Stephan Ripke, Jian Yang, Nick Patterson, Mark J. Daly, Alkes L. Price, Benjamin M. Neale, and Schizophrenia Working Group of the Psychiatric Genomics Consortium. Ld score regression distinguishes confounding from polygenicity in genome-wide association studies. Nature Genetics, 47(3):291–295, Mar 2015.

42. Wouter van Rheenen, Wouter J. Peyrot, Andrew J. Schork, S. Hong Lee, and Naomi R. Wray. Genetic correlations of polygenic disease traits: from theory to practice. Nature Reviews Genetics, 20(10):567–581, Oct 2019.

43. Signe M. Sørensen, Henrik C. Schønheyder, and Henrik Nielsen. The role of imaging of the urinary tract in patients with urosepsis. International Journal of Infectious Diseases, 17(5):e299–e303, May 2013.

44. Ruowang Li, Yong Chen, Marylyn D. Ritchie, and Jason H. Moore. Electronic health records and polygenic risk scores for predicting disease risk. Nature Reviews Genetics, 21(8):493– 502, Aug 2020.

45. Grimur Hjorleifsson Eldjarn, Egil Ferkingstad, Sigrun H. Lund, Hannes Helgason, Olafur Th. Magnusson, Kristbjorg Gunnarsdottir, Thorunn A. Olafsdottir, Bjarni V. Halldorsson, Pall I. Olason, Florian Zink, Sigurjon A. Gudjonsson, Gardar Sveinbjornsson, Magnus I. Magnusson, Agnar Helgason, Asmundur Oddsson, Gisli H. Halldorsson, Magnus K. Magnusson, Saedis Saevarsdottir, Thjodbjorg Eiriksdottir, Gisli Masson, Hreinn Stefansson, Ingileif Jonsdottir, Hilma Holm, Thorunn Rafnar, Pall Melsted, Jona Saemundsdottir, Gudmundur L. Norddahl, Gudmar Thorleifsson, Magnus O. Ulfarsson, Daniel F. Gudbjartsson, Unnur Thorsteinsdottir, Patrick Sulem, and Kari Stefansson. Large-scale plasma proteomics comparisons through genetics and disease associations. Nature, 622(7982):348–358, Oct 2023.

46. Clare Bycroft, Colin Freeman, Desislava Petkova, Gavin Band, Lloyd T. Elliott, Kevin Sharp, Allan Motyer, Damjan Vukcevic, Olivier Delaneau, Jared O’Connell, Adrian Cortes, Samantha Welsh, Alan Young, Mark Effingham, Gil McVean, Stephen Leslie, Naomi Allen, Peter Donnelly, and Jonathan Marchini. The uk biobank resource with deep phenotyping and genomic data. Nature (London), 562(7726):203–209, 2018.

47. Paul Elliott and on behalf of UK Biobank Peakman, Tim C. The uk biobank sample handling and storage protocol for the collection, processing and archiving of human blood and urine. International Journal of Epidemiology, 37(2):234–244, 04 2008.

48. AF Agarap. Deep learning using rectified linear units (relu). arXiv preprint 1803.08375, 2018.

49. Lei Jimmy Ba, Jamie Ryan Kiros, and Geoffrey E. Hinton. Layer normalization. CoRR, abs/1607.06450, 2016.

50. Yingyao Zhou, Bin Zhou, Lars Pache, Max Chang, Alireza Hadj Khodabakhshi, Olga Tanase-ichuk, Christopher Benner, and Sumit K Chanda. Metascape provides a biologist-oriented resource for the analysis of systems-level datasets. Nature communications, 10(1):1523, 2019.

