## Supplementary figures and images for "A multistage, multitask transformer-based framework for multi-disease diagnosis and prediction using personal proteomes"

### Supplementary Fig. 1

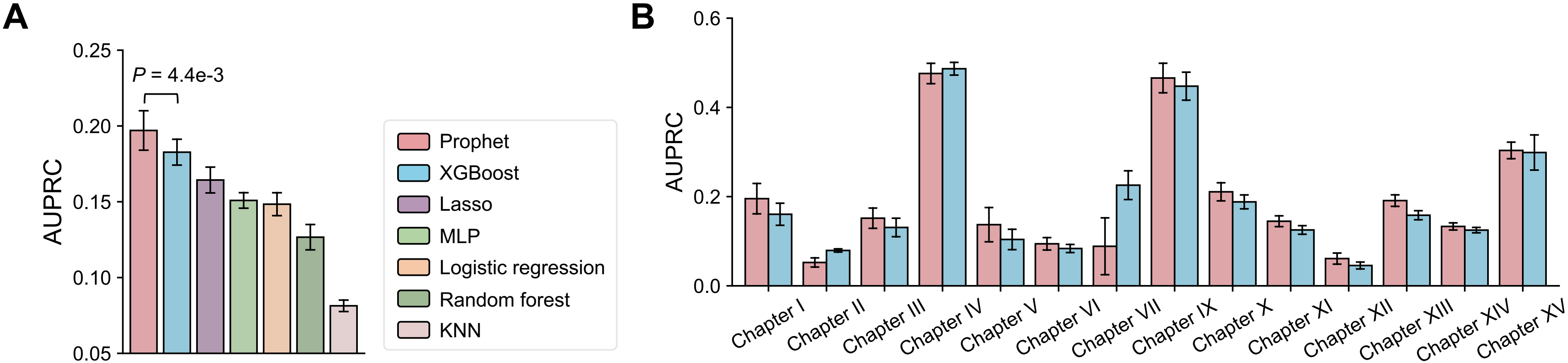

### Supplementary Fig. 2

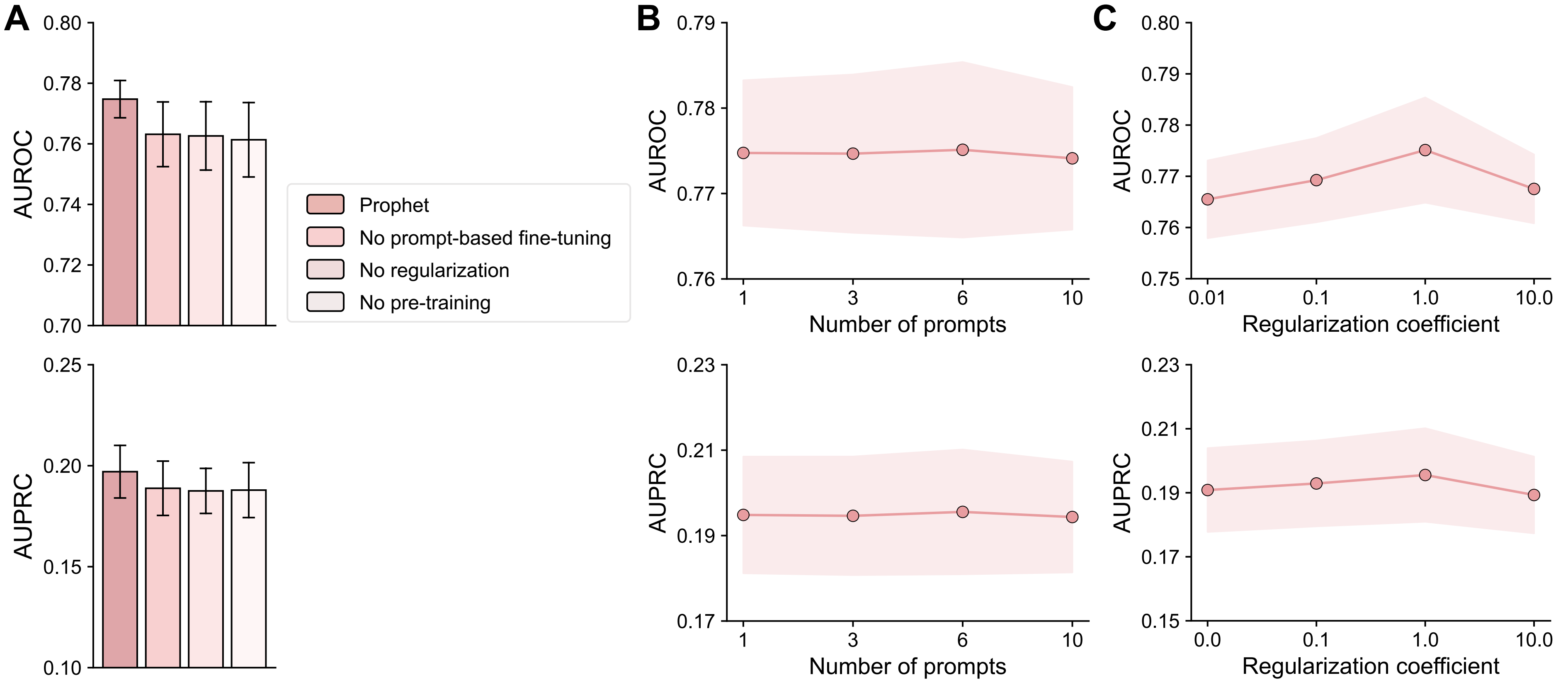

### Supplementary Fig. 3

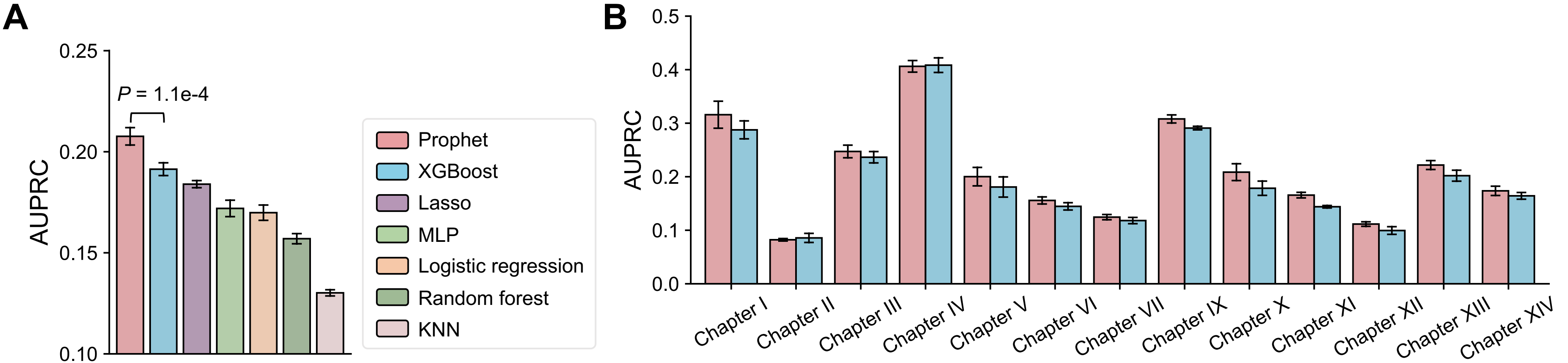

### Supplementary Fig. 4

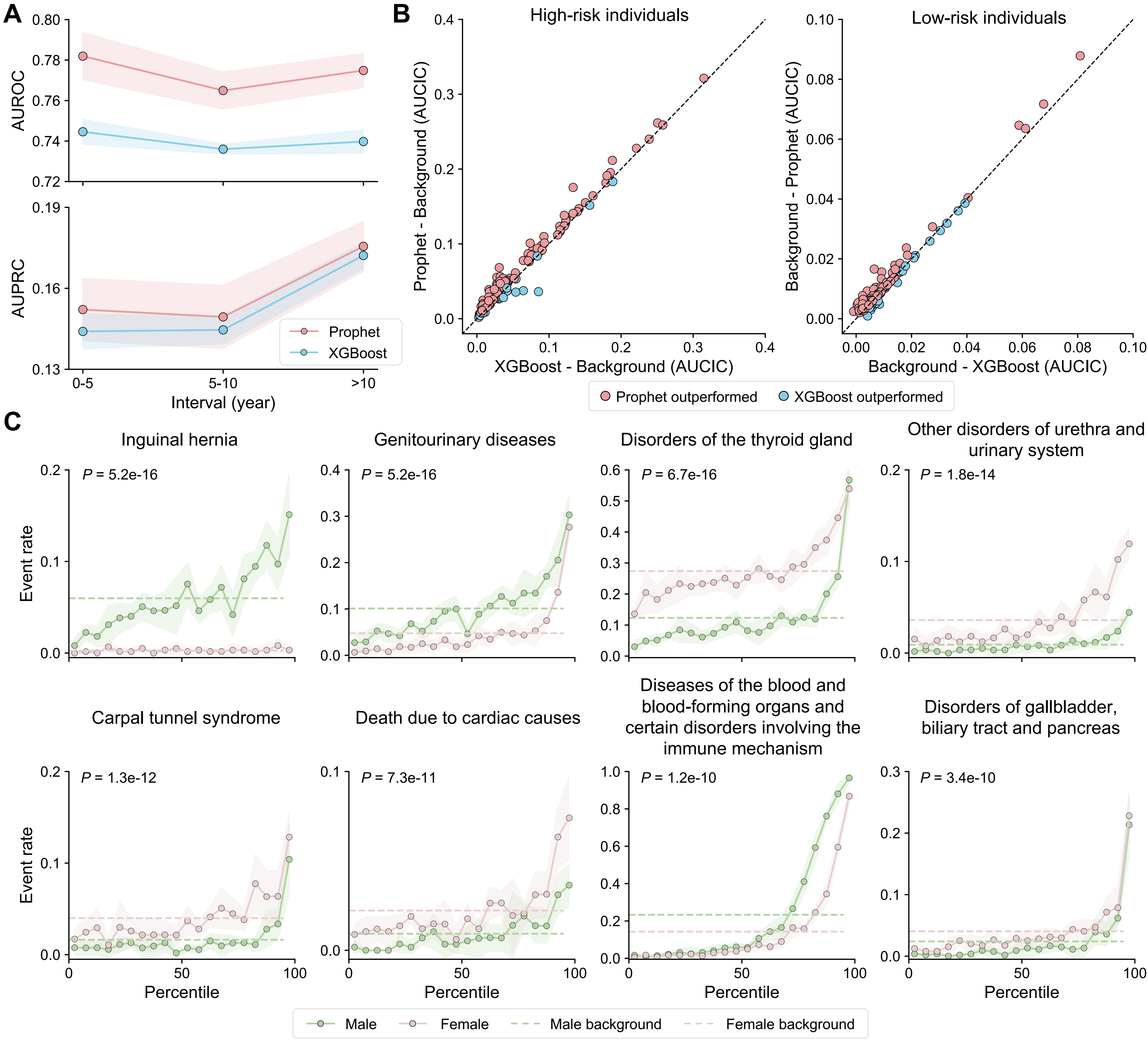

### Supplementary Fig. 5

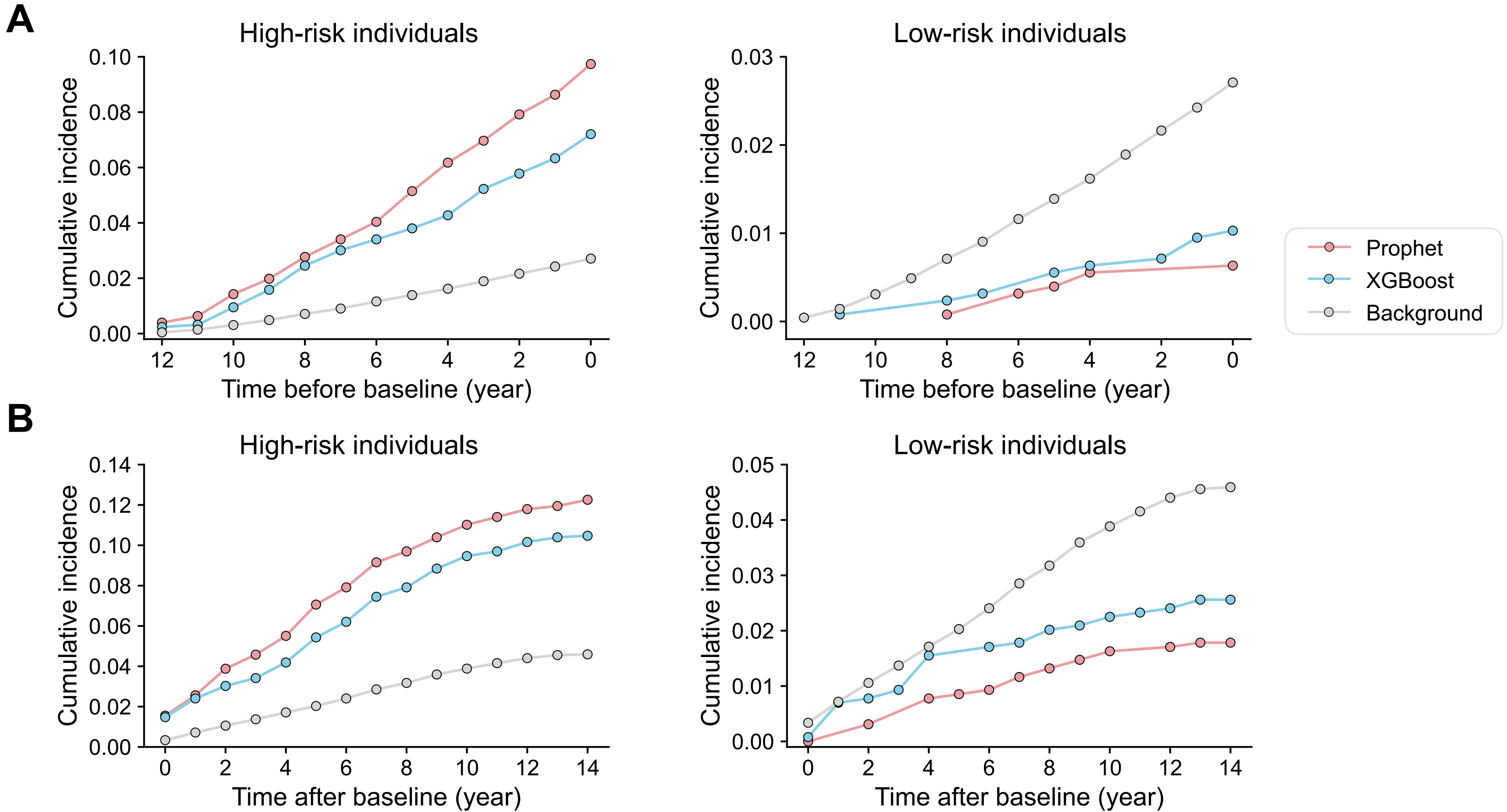

### Supplementary Fig. 6

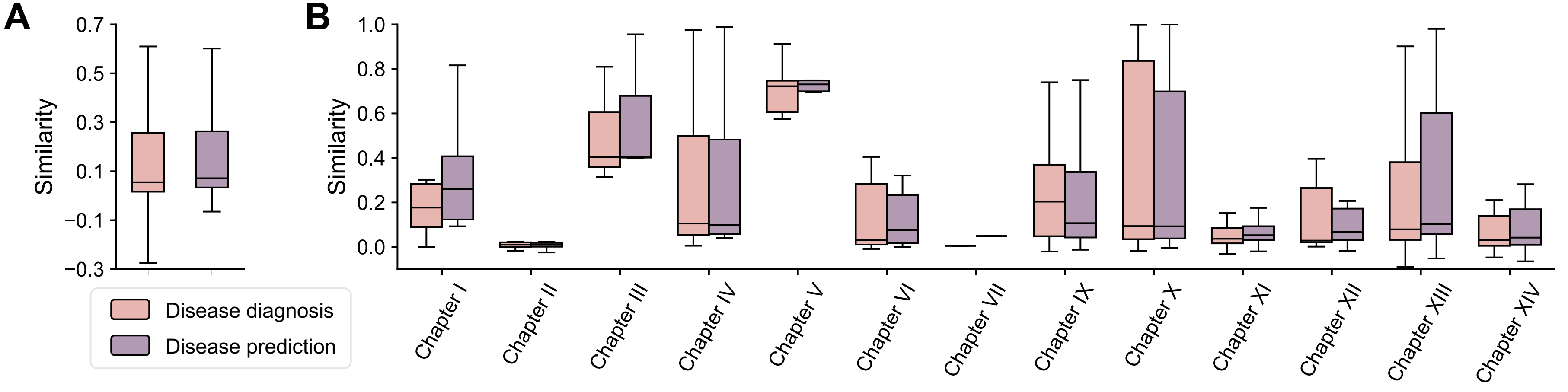
